# Analytical validation and proof-of-concept prognostic evaluation of plasma-membrane TRPV4 localization in ductal carcinoma in situ: a retrospective case-control study

**DOI:** 10.1101/2025.11.04.25339245

**Authors:** Christopher Chan, Elham Arbzadeh, Shabnam Samankan, Patricia Latham, Arnold Schwartz, Rachel Du, Inhee Chung

## Abstract

**Purpose:** Ductal carcinoma in situ (DCIS) is a non-invasive breast neoplasm frequently overtreated because biomarkers of invasive progression are lacking. We previously identified a pro-invasive mechanotransduction pathway in which ductal cell crowding inactivates plasma-membrane (PM) transient receptor potential vanilloid 4 (TRPV4), driving compensatory PM relocalization of intracellular inactive TRPV4 that marks pathway-engaged cells. Here, we evaluated whether PM-associated TRPV4 immunohistochemical staining on routine formalin-fixed paraffin-embedded DCIS sections is associated with subsequent invasive progression and assessed analytical validation of the antibody and scoring method.

**Methods:** We performed a retrospective single-institution case-control study of 44 patients with pure DCIS: 24 who subsequently developed invasive ductal carcinoma and 20 who remained free of invasive progression for ≥5 years. TRPV4 immunohistochemistry was scored for PM-TRPV4 on 225 pathology-annotated regions of interest (ROI) by three board-certified pathologists using a prespecified five-level rubric dichotomized for primary analyses as PM-positive or PM-negative. Orthogonal validation used a second human immunohistochemistry-validated TRPV4 antibody in representative ROI and cell-line models.

**Results:** Orthogonal validation confirmed concordant compartmental TRPV4 distributions and mechanosensitive PM-TRPV4 relocalization in cell-line models. PM-TRPV4 scoring was highly reproducible (weighted Fleiss’ κ=0.823, 95% CI 0.777-0.863). PM-TRPV4 positivity was more frequent in progressors than non-progressors (22/24 vs 13/20), linked to shorter invasive progression-free survival (log-rank p=0.040), and was independently prognostic after adjustment for grade and estrogen receptor status (adjusted HR 3.77, 95% CI 1.01-14.12; p=0.049).

**Conclusions:** These findings support PM-TRPV4 as a mechanism-informed, pathology-interpretable biomarker for DCIS risk stratification, with reproducible scoring and a preliminary prognostic signal warranting multi-institutional validation.

## Introduction

Ductal carcinoma in situ (DCIS) is a non-invasive breast neoplasm that challenges clinical decision-making because the risk of progression to invasive ductal carcinoma (IDC) remains difficult to predict for the more than 60,000 women who are diagnosed annually in the United States (1–4). This uncertainty contributes to overtreatment of indolent lesions and insufficient escalation for high-risk disease. Histologic grading, based on nuclear morphology, architecture, and necrosis, provides some prognostic information but shows only moderate interobserver reproducibility (κ ∼0.4-0.6) and limited accuracy for predicting progression (5–14). The problem is most acute in lower-grade DCIS (intermediate+low), which represents ∼60% of cases (12, 15, 16), where morphology alone offers limited guidance, because some lesions still invade (17, 18). Biomarkers that discriminate risk within this large subgroup could enable more personalized management.

Emerging evidence points beyond morphology to microenvironmental and patient-level biology in the progression of DCIS to IDC. Extracellular matrix remodeling has been linked to progression (19, 20); molecular programs associate with outcomes (21); and biomarker profiles predict IDC events after DCIS diagnosis (17, 18). Active-surveillance trials (COMET, LORD, LORIS) demonstrate feasibility in carefully selected low-risk DCIS, underscoring the need for robust biomarkers to support de-escalation where appropriate (22–25). Genomic tools such as DCISionRT can inform radiotherapy decisions (26) but require specialized assays and may be less accessible, motivating complementary biomarkers measurable on routine FFPE sections.

Guided by recent findings from our group, we hypothesized that mechanical stress responses provide a biological basis for DCIS progression risk (27). In confined ductal spaces, epithelial cell crowding inhibits calcium ion channels such as transient receptor potential vanilloid 4 (TRPV4) in the plasma membrane, lowers intracellular calcium, remodels the cortical cytoskeleton, and alters cell mechanics, which promotes pro-invasive behaviors (27–31). During this process, inactive intracellular TRPV4 undergoes compensatory relocalization to the plasma membrane, where it is poised for reactivation by subsequent mechanical cues (27). Importantly, TRPV4 biology is subcellularly compartmentalized, and in our prior work (27) as well as the present study, TRPV4 signal is observed in both nuclear and plasma-membrane compartments. This PM-accumulated inactive TRPV4 represents a primed pool, spatially positioned to sense and respond to future mechanical inputs, potentially lowering the threshold for invasive transition. Recent studies (32, 33) have identified a functional nuclear localization signal in TRPV4 and demonstrated regulated trafficking between nuclear and plasma-membrane compartments, supporting nuclear TRPV4 as a biologically regulated localization state rather than inherently nonspecific signal. Critically, these findings support the PM-associated TRPV4 staining pattern as a specific, mechanistically grounded readout worth clinical testing. We therefore asked whether this PM-associated pattern can be reproducibly scored on routine FFPE sections. This mechanosensing-linked PM relocalization, distinct from total TRPV4 abundance or non-membranous localization (27), provides a functional readout of pro-invasive pathway engagement that may complement morphology-based grading for risk stratification in DCIS. By capturing a stress-responsive cellular state rather than static histologic features alone, PM-TRPV4 may improve prognostic discrimination, particularly in lower-grade disease, where clinical decision-making remains especially challenging (12, 15, 16).

In this retrospective case-control study, our objectives were to (i) strengthen analytical confidence in TRPV4 staining interpretation on human FFPE material, including orthogonal antibody-validation data; (ii) assess the association between PM-TRPV4 status and time to invasive progression; (iii) determine whether PM-TRPV4 provides prognostic information independent of histologic grade and estrogen receptor (ER) status; and (iv) evaluate performance across grade strata, with emphasis on lower-grade DCIS, where improved clinical utility is most needed. Because PM-TRPV4 was hypothesized to reflect a biologically relevant lesion state rather than laterality alone, invasive events were analyzed irrespective of laterality (ipsilateral or contralateral), and prespecified sensitivity analyses evaluated potential confounding by treatment selection and event laterality.

## Methods

### Study design and ethical oversight

We conducted a retrospective case-control study at George Washington University comparing patients with pure ductal carcinoma in situ (DCIS) who subsequently developed invasive ductal carcinoma (IDC) with patients who remained free of invasive progression (non-progressors). Group assignment to case vs non-progressor groups was based on clinical outcome rather than randomization, consistent with a retrospective case–control design. Sample size was determined by the availability of eligible cases with adequate tissue quality and documented outcomes from the biorepository. No formal a priori power calculation was performed, as this was an exploratory biomarker discovery study designed to generate hypotheses for validation in larger cohorts. The study was approved by the George Washington University Institutional Review Board (protocol NCR203065) with a waiver of informed consent for archived tissue and de-identified clinical data.

### Patient cohort and tissue procurement

Formalin-fixed paraffin-embedded (FFPE) tissue blocks and associated de-identified clinical data were obtained from the Cooperative Human Tissue Network (CHTN), a National Cancer Institute-supported biorepository that prospectively collects specimens from multiple academic medical centers. Samples represented DCIS diagnoses from 2005-2021. From 51 screened subjects, 44 met all criteria and comprised the analytic cohort. FFPE blocks were stored under standard archival conditions at room temperature at CHTN prior to shipment; precise fixation duration and cumulative storage time were not uniformly available for all cases, a limitation inherent to biorepository material (**Fig. 1**, **Supplementary Table S1**).

**Figure 1.**
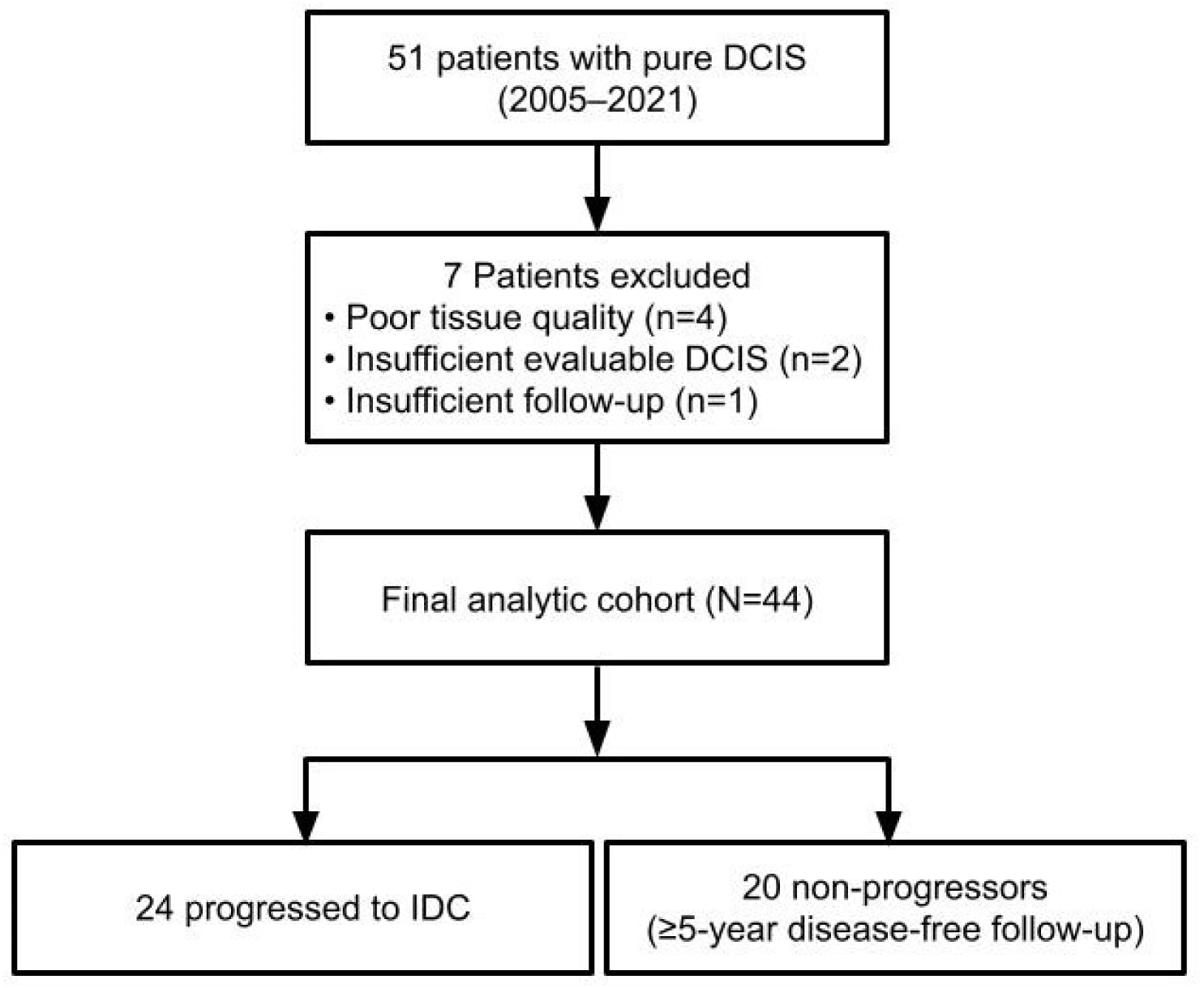
Patient selection and study flow diagram. Flow diagram showing patient selection for DCIS progression analysis. From an initial cohort of 51 retrospective patients with FFPE tissue samples and pure ductal carcinoma in situ, 7 cases were excluded because of inadequate tissue quality (n=4), insufficient evaluable DCIS regions (n=2), or short follow-up (n=1), yielding a final analytical cohort of 44 patients. Twenty-four patients (54.5%) experienced subsequent invasive progression during follow-up, while 20 patients (45.5%) remained without invasive progression during a minimum of 5 years of surveillance.

### Inclusion criteria

i. histologically confirmed pure DCIS at index diagnosis; (ii) adequate FFPE tissue for immunohistochemistry (IHC); (iii) clinical follow-up sufficient to document invasive outcomes or free from invasive progression; (iv) complete pathology records including histologic grade.

### Exclusion criteria

i. concurrent invasive carcinoma at index diagnosis; (ii) prior invasive breast cancer; (iii) insufficient tissue quantity/quality for biomarker assessment; (iv) inadequate evaluable DCIS regions for scoring; (v) insufficient follow-up to confirm non-progression (i.e., <5 years free of invasive progression among those without an invasive event).

Seven subjects were excluded: insufficient tissue quality (n=4), inadequate evaluable DCIS regions for scoring (n=2), and insufficient follow-up (n=1). Progressors were included regardless of surveillance duration; the requirement for ≥5 year free of invasive progression applied only to non-progressors.

### Case-control definitions and outcome assessment

Cases (n=24): patients with pure DCIS at diagnosis who subsequently developed histologically confirmed IDC during follow-up (ipsilateral or contralateral breast; chest wall after mastectomy; or regional lymph nodes). Because PM-TRPV4 was hypothesized to report a patient-level mechanosensing state, invasive events were analyzed irrespective of laterality; laterality was evaluated in sensitivity analyses.

Non-progressors (n=20): patients with pure DCIS who remained free of invasive progression for ≥5 years of documented surveillance, based on clinical examination, imaging, and absence of histologic invasion on any subsequent biopsies.

The primary endpoint was time from index DCIS diagnosis to the first invasive event as defined above; patients without an event were censored at last contact free of invasive progression.

### Clinical data abstraction

All patients in this study were female. Clinical and pathologic variables abstracted from records included age, self-reported race/ethnicity, histologic grade at diagnosis, estrogen receptor (ER) and progesterone receptor (PR) status, surgical treatment (breast-conserving surgery vs mastectomy), receipt of adjuvant radiation, and comorbidities (cardiovascular disease, metabolic disease, smoking history). ER was available for 42/44 patients; PR was sparsely recorded and not used in adjusted models. Median age at diagnosis was 62 years (range, 39-79). Histologic grade was called as low, intermediate, or high by standard criteria and was also collapsed a priori to a binary variable: high grade (HG) vs lower grade (intermediate+low). Treatment decisions were made at the time of diagnosis according to institutional standard of care and patient preference; biomarker status was unknown at the time of treatment.

### Immunohistochemistry for TRPV4 and image acquisition

IHC was performed on 4-μm FFPE sections using a Ventana Discovery Ultra platform. Sections were deparaffinized with EZ Prep, subjected to heat-induced epitope retrieval with Cell Conditioner 1 (CC1; Tris-EDTA, pH 8.0) for 64 min at 95 °C, and treated with endogenous peroxidase block (CM1) for 8 min. The primary antibody was anti-TRPV4 (Abcam Cat# ab39260, RRID:AB_1143677; 1:40; 60 min at room temperature), validated by knockdown (KD) in our prior mechanistic study (27). Detection used DISCOVERY OmniMap anti-rabbit HRP (Roche Cat# 760-4311, RRID:AB_2811043) and ChromoMap DAB (Roche Cat# 05266645001), followed by hematoxylin counterstaining, bluing, dehydration, and permanent mounting. Each run included human duodenum as a positive-control tissue and primary-antibody-omitted sections as negative controls; representative images are shown in Supplementary Fig. S1. Whole-slide images were acquired on a Hamamatsu NanoZoomer at 20× (0.46 μm/pixel). At the time of study initiation, no commercially available anti-TRPV4 antibody carried formal human FFPE IHC validation; ab39260 was selected for its published application in human tissue and superior signal intensity across the full five-level scoring rubric (27). To assess whether the scored staining pattern reflects true compartmental biology rather than reagent-specific signal, we performed a retrospective bridging comparison using a second, knockdown/knockout (KD/KO)-validated antibody (MilliporeSigma Cat# ZRB3155, clone 1G10-L2; 1:30) on matched DCIS sections. Line-scan analyses across five paired ROIs (10 cells per ROI) compared PM, nuclear, cytosolic, whole-cell, and PM:nuclear signal distributions between antibodies (**Supplementary Fig. S2**, **Supplementary Table S2,S3**, **Fig. 2-3**). Both antibodies showed closely aligned compartmental profiles, confirming that the localization pattern, not absolute DAB intensity, is the reproducible feature underlying PM-TRPV4 scoring. All pathologist scoring and primary outcome analyses were performed using sections stained with Abcam ab39260; MilliporeSigma ZRB3155 was used only for orthogonal bridging validation on matched tissue and in cell-line models.

**Figure 2.**
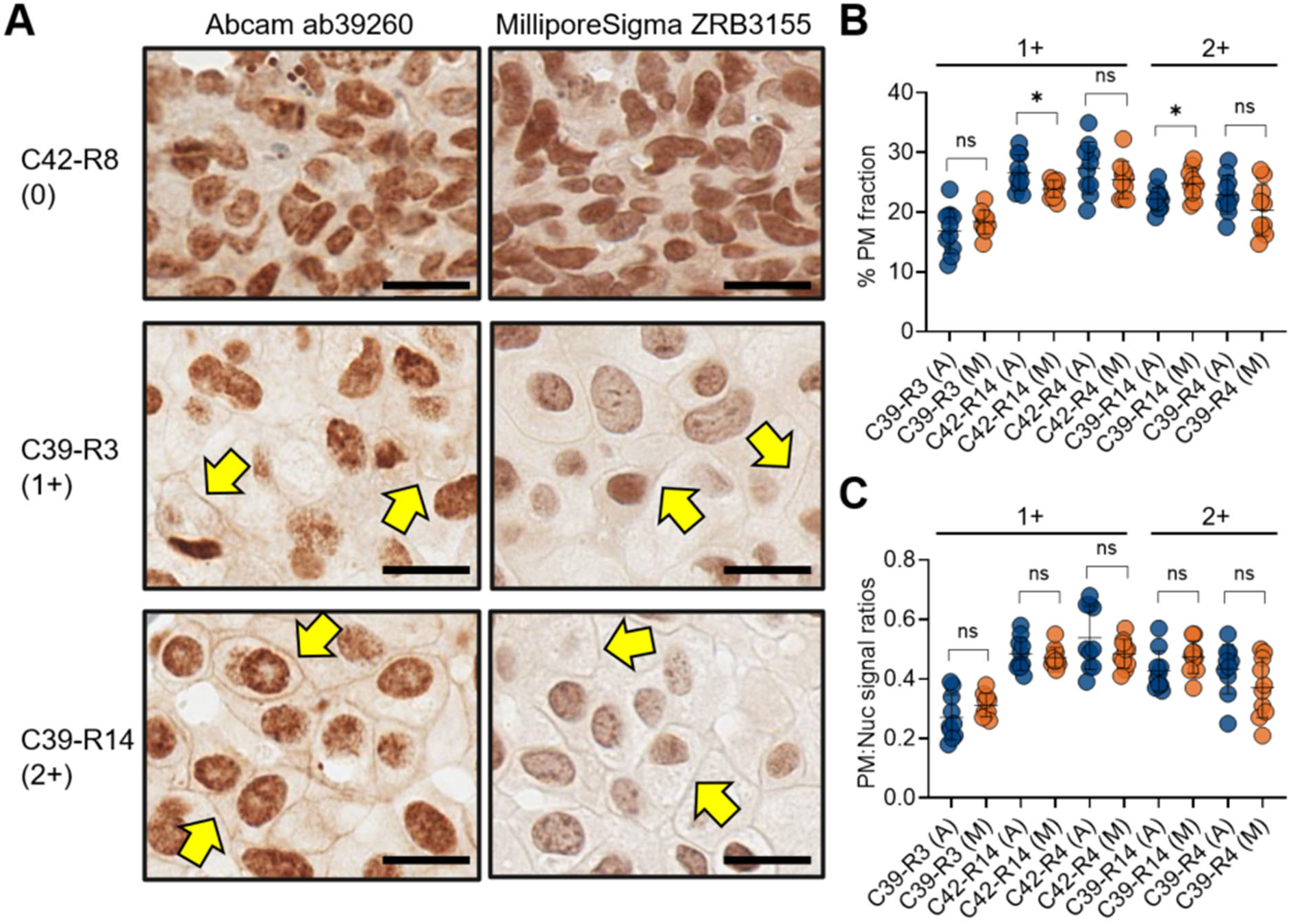
Orthogonal antibody validation supports preservation of TRPV4 compartmental localization patterns in representative matched ROIs. **(A)** Representative matched ROIs stained with the primary study antibody (Abcam ab39260) or the orthogonal validation antibody (MilliporeSigma ZRB3155) show similar compartmental TRPV4 staining patterns, including preservation of a discernible plasma-membrane rim in selected cells (yellow arrows). Scale bars: 100 µm. **(B)** Cell-level comparison of PM fraction across matched ROIs from 1+ and 2+ groups. **(C)** Cell-level comparison of PM:nuclear signal ratio across the same ROIs. Blue points indicate Abcam ab39260 (A) and orange points indicate MilliporeSigma ZRB3155 (M). Each point represents one analyzed cell; horizontal bars indicate mean ± SD. Statistical comparisons were performed between antibodies within each matched ROI; ns, not significant; *, p < 0.05.

**Figure 3.**
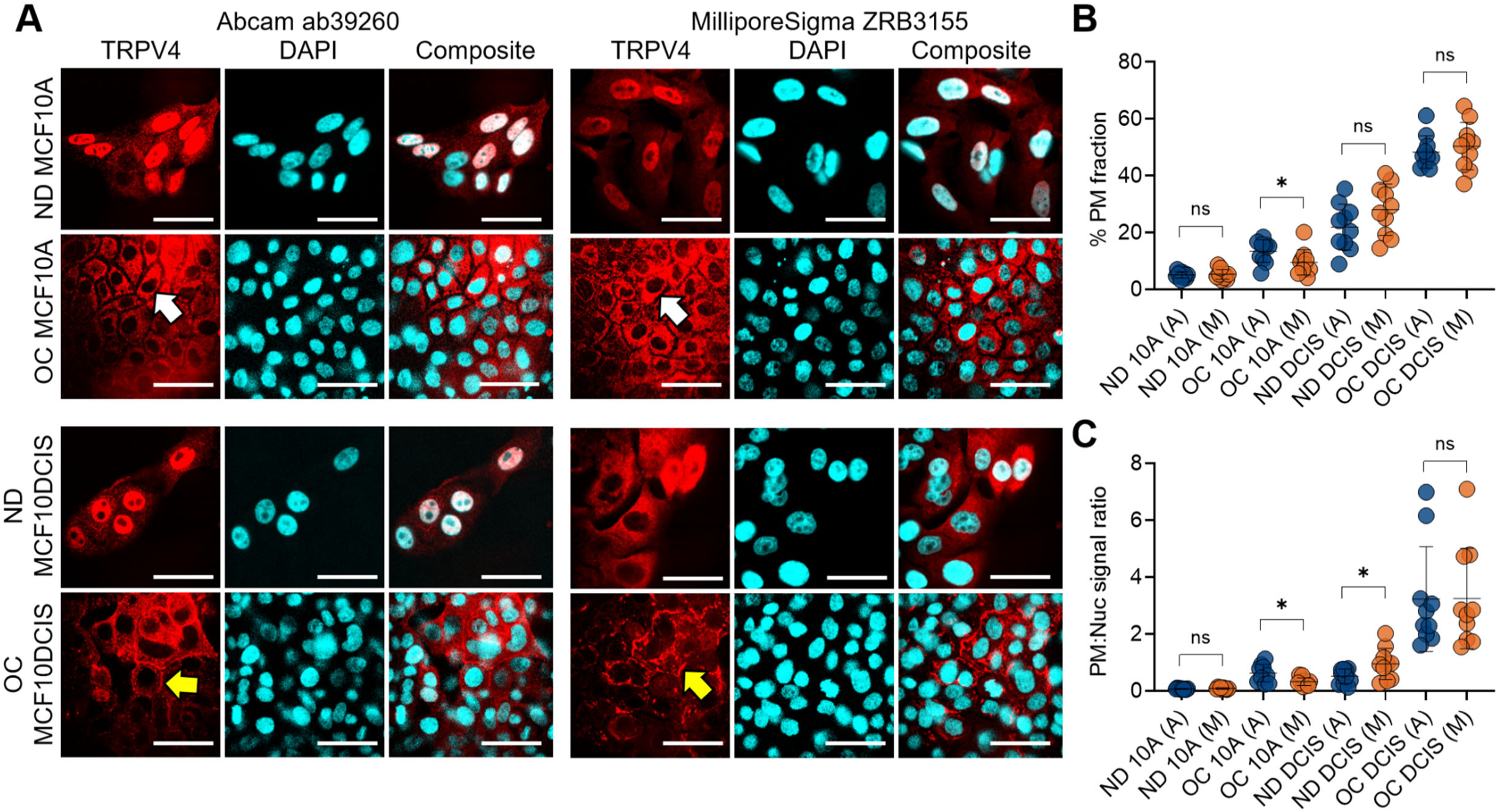
An orthogonal validation TRPV4 antibody reproduces mechanosensing-linked PM relocalization in cell models. **(A)** Representative immunofluorescence images comparing TRPV4 localization detected with Abcam ab39260 and MilliporeSigma ZRB3155 in MCF10A and MCF10DCIS.com cells under normal-density (ND) and overconfluent (OC) conditions. TRPV4 is shown in red, nuclei in cyan, and merged images at right. In OC MCF10A cells, white arrows indicate the absence of discernible PM TRPV4 enrichment under crowding stress, whereas in OC MCF10DCIS.com cells, yellow arrows indicate representative cells with discernible PM rim-associated TRPV4 signal under crowding stress. **(B)** Cell-level comparison of PM fraction between antibodies across the indicated conditions. **(C)** Cell-level comparison of PM:nuclear signal ratio across the same conditions. Blue points indicate Abcam ab39260 (A) and orange points indicate MilliporeSigma ZRB3155 (M). Each point represents one analyzed cell; horizontal bars indicate mean ± SD. Statistical comparisons were performed between antibodies within the same conditions; ns, not significant; p < 0.05. Scale bars, 50 µm.

### PM-TRPV4 scoring rubric and blinding

PM-TRPV4 relocalization was scored on a five-level ordinal scale incorporating (1) % DCIS epithelial cells with PM TRPV4 staining, (2) circumferential PM coverage, and (3) staining intensity:

(negative): <5% cells; incomplete/absent membrane; faint/absent intensity.

0-1 + (trace): ≥5% and <10% cells; incomplete membrane; faint intensity (below the 1+ threshold).

1+ (low-positive): ≥10% cells; partial to complete membrane; weak to focally moderate intensity.

2+ (moderate-positive): ≥10% cells with ≥50% circumferential coverage; moderate intensity.

3+ (strong-positive): ≥10% cells with ≥50% circumferential coverage; strong intensity.

Three board-certified breast pathologists independently scored 225 image fields from 44 patients, blinded to outcomes, clinical covariates, one another’s scores, and grade assignments. Only epithelial membrane staining contributed to the PM-TRPV4 score; nuclear staining intensity was not incorporated into scoring of PM positivity. For primary analyses, scores were dichotomized a priori:

PM-TRPV4 positive (PM+): if any rater assigned ≥1+, or if all three raters unanimously assigned 0-1+ (trace).

PM-TRPV4 negative (PM-): if all three raters assigned 0, or any mixture of 0 and 0-1+ (discordant trace signal).

This rule prioritizes definite PM relocalization while conservatively classifying ambiguous faint/discordant membrane signal as negative; unanimous trace was treated as biologically positive. Diffuse nuclear or cytoplasmic staining without a discernible membrane rim was not sufficient for PM positivity. The five-level rubric was used to capture the full spectrum of membrane-associated staining, whereas dichotomization was prespecified for primary analyses to maximize interpretability and analytical robustness in this modest-sized cohort.

### Interobserver agreement

Agreement on the five-level scale was quantified using linearly weighted Fleiss’ κ and unweighted Fleiss’ κ; pairwise agreement was summarized with weighted Cohen’s κ. 95% confidence intervals (CIs) were obtained from asymptotic variance; exact agreement (3/3 concordance and 2/3 majority) was summarized as n (%).

## Statistical analysis

All tests were two-sided with α=0.05. Analyses used MATLAB R2022a (MathWorks, RRID: SCR_001622) and Python 3.12 (python-bx RRID: SCR_024202, SciPy 1.16.2 RRID: SCR_008058, NumPy: 2.0.2 RRID: SCR_008633). Continuous variables are summarized as median or mean ± standard deviation (SD) or standard error (SE); categorical variables as counts (%).

### Time-to-event analyses

Invasive progression-free survival was estimated by Kaplan-Meier with 95% CIs by Greenwood and compared using the log-rank test. Cox proportional-hazards models reported hazard ratios (HRs) with 95% Wald CIs and Wald p-values. Ties were handled using the Efron method. Proportional-hazards assumptions were assessed for each covariate using Schoenfeld residual tests.

### Prespecified models

Univariable: PM-TRPV4 (primary predictor).

Univariable: histologic grade (high grade vs lower grades).

Multivariable: PM-TRPV4 + histologic grade.

Multivariable: PM-TRPV4 + ER status.

Multivariable (fully adjusted): PM-TRPV4 + histologic grade + ER status.

### Case-control (2×2) association analyses

To complement survival analyses, Fisher’s exact tests (two-sided) were applied to 2×2 tables with odds ratios (ORs) and 95% CIs computed by the Woolf logit method with Haldane-Anscombe correction for zero cells. A prespecified one-sided Fisher test was additionally performed for the directional hypothesis that PM-TRPV4 relocalization increases the risk of invasive progression. Because case-control sampling fixes the case:control ratio, absolute risk metrics (PPV/NPV/accuracy) are not estimable and were not reported. Exploratory sensitivity analyses of treatment selection and event laterality used the same approach and are provided in **Supplementary Tables S5-S9**. Variables collected but not included in adjusted models were: PR status (recorded in fewer than 10% of cases; excluded from all models) and comorbidities (cardiovascular disease, metabolic disease, smoking history; evaluated in univariable sensitivity analyses in **Supplementary Tables S5-S9**, none significant). ER status was missing for 2/44 patients; these patients were excluded from ER-adjusted Cox models (n=42 for ER-adjusted analyses) but retained in all other analyses. No imputation was performed; all other variables had complete data. No correction for multiple comparisons was applied. Subgroup analyses by histologic grade and all sensitivity analyses were pre-specified but exploratory; findings from these analyses should be interpreted as hypothesis-generating rather than confirmatory.

## Results

### Patient characteristics

From 51 retrospectively identified patients with pure DCIS, 44 were included after exclusions for poor tissue quality (n=4), insufficient evaluable DCIS regions (n=2), or inadequate follow-up (n=1) (**Fig. 1**; **Table 1**). Median age at diagnosis was 62 years (range, 39-79). Race: White 30/44 (68.2%), Black 10/44 (22.7%), Asian 1/44 (2.3%), other/unknown 3/44 (6.8%). Nuclear grade: high 18/44 (40.9%), intermediate 14/44 (31.8%), low 12/44 (27.3%). ER status was available for 42/44 (ER+ 35/42, 83.3%). Primary surgery: lumpectomy 32/44 (72.7%), mastectomy 11/44 (25.0%) (other/unknown 1/44, 2.3%). By design, 24 were progressors and 20 non-progressors with ≥ 5 years of invasive progression-free surveillance (**Supplementary Table S1**).

**Table 1.** Patient demographics and clinical characteristics. Baseline characteristics of patients with pure DCIS (N=44). Data presented as n (%) for categorical variables and median (range) for continuous variables. DCIS grade was determined by board-certified pathologists using standard criteria including nuclear morphology and architectural patterns. Treatment decisions were made according to institutional guidelines and patient preference. *ER status was available for 42 of 44 patients.

| Characteristic | Total (N = 44) |
| --- | --- |
| Ages (39-79 years) |  |
| Age, median | 62 |
| Race |  |
| White | 30 (68.2%) |
| Black | 10 (22.7%) |
| Asian | 1 (2.3%) |
| Other | 3 (6.8%) |
| DCIS Grade |  |
| High | 18 (40.9%) |
| Intermediate | 14 (31.8%) |
| Low | 12 (27.3%) |
| ER Status* (n=42) |  |
| Positive | 35 (83.3%) |
| Negative | 7 (16.7%) |
| Treatment |  |
| Lumpectomy | 32 (72.7%) |
| Mastectomy | 11 (25.0%) |
| Radiation | 23 (52.3%) |
| *\*n = 42, after excluding cases with unknown ER status* |  |

### PM-TRPV4 scoring and inter-observer agreement

On each staining run, human duodenum served as a positive-tissue control confirming expected enterocyte TRPV4 expression, and omission of the primary antibody confirmed assay specificity (**Supplementary Fig. S1**). We then performed orthogonal bridging validation with a second antibody to confirm that the scored staining pattern reflected reproducible compartment-specific TRPV4 localization on human FFPE DCIS tissue before outcome analysis. Across five matched ROIs, the Abcam and MilliporeSigma antibodies showed closely concordant compartmental distributions: PM fractions of 0.232 versus 0.226, nuclear fractions of 0.552 versus 0.542, and PM:nuclear ratios of 0.431 versus 0.423, corresponding to only 1.8–2.6% deviation across compartment-level metrics (all <3%; **Supplementary Table S2**). Nuclear signal remained the dominant compartment in every ROI pair, while a discrete plasma-membrane rim signal was detectable with both antibodies (**Fig. 2**). Absolute staining intensity was approximately 1.8-fold higher with Abcam (background-subtracted single-cell intensity, 64.1 vs 34.9 arbitrary units; see **Supplementary Fig. S2C,D** for single-cell intensity values); however, both antibodies showed concordant subcellular distribution patterns, confirming that compartment balance rather than absolute intensity is the reproducible feature underlying PM-TRPV4 scoring (**Supplementary Fig. S2**; **Fig. 2**). Together, these findings support nuclear TRPV4 as a genuine subcellular localization state in DCIS cells and show that the PM-relocalized pool is specifically detectable and distinguishable from non-membranous signal with either antibody.

To further confirm that the orthogonally validated staining pattern reflects the mechanosensing-induced PM relocalization signal, we examined them by immunofluorescence in MCF10DCIS.com cell-line models, the model system in which the crowding-induced pro-invasive mechanotransduction mechanism was originally identified (27), under overconfluent (OC) versus normal density (ND) conditions. Under overcrowded conditions, both antibodies showed PM-dominant distributions: mean PM:nuclear ratios of 3.23 ± 1.85 (Abcam) and 3.25 ± 1.77 (MilliporeSigma), with less than 1% deviation between antibodies (**Fig. 3**). Under normal density conditions, both antibodies showed nuclear-dominant distributions (Abcam 0.51 ± 0.25; MilliporeSigma 0.95 ± 0.55); these values were not significantly different between antibodies, consistent with low PM signal and staining heterogeneity at baseline. The direction of crowding-induced PM relocalization was concordant across antibodies (**Fig. 3**). This relocalization was absent in non-mechanosensing MCF10A cells (27), confirming that the PM-TRPV4 signal is specific to the pro-invasive mechanosensing context rather than a generic membrane staining artifact.

PM-TRPV4 localization was then assessed on whole-slide images using a prespecified five-level rubric (0, 0-1+, 1+, 2+, 3+; **Fig. 4A-E**), based on the proportion of DCIS cells exhibiting membranous staining, circumferential PM coverage, and staining intensity. Three board-certified pathologists independently scored 225 image fields from 44 patients, blinded to clinical outcomes. For primary analyses, scores were dichotomized a priori: PM-TRPV4 positive if ≥ 1 rater assigned ≥ 1+ or if all three raters unanimously assigned 0-1+ (indicating consistent faint membranous signal); PM-TRPV4 negative if all three raters assigned 0 or if scores were any mixture of 0 and 0-1+ (discordant faint signal). The five-level rubric was used to capture the full spectrum of membrane-associated staining, whereas dichotomization was prespecified for primary analyses to maximize interpretability and analytical robustness in this modest-sized cohort while conservatively classifying ambiguous cases as negative (see **Supplementary Data S1** for score-driving ROI images for each case).

**Figure 4.**
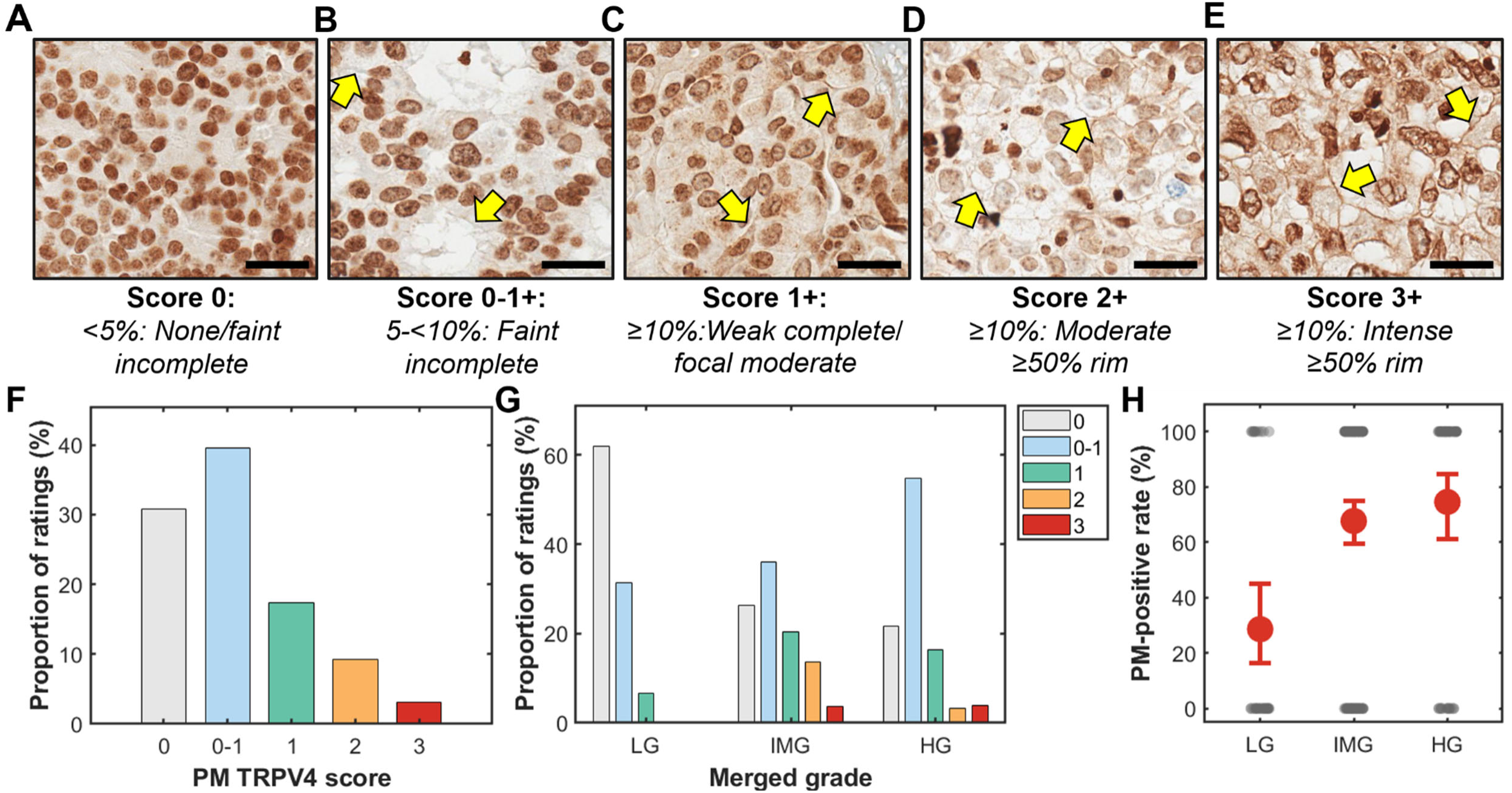
PM-TRPV4 Scoring Rubric, Score Distributions, and Association with Grade. **(A–E)** Representative whole-slide image fields illustrating the 5-level PM-TRPV4 scoring rubric applied to DCIS epithelial cells. Yellow arrows indicate cells with PM-associated TRPV4 staining. **(A)** Score 0: No staining, or incomplete and faint PM staining in <5% of DCIS cells. **(B)** Score 0-1+: Incomplete and faint PM staining in ≥5% and <10% of DCIS cells. **(C)** Score 1+: Weak complete to focally moderate PM staining in ≥10% of DCIS cells, with PM coverage ranging from partial to complete. **(D)** Score 2+: Moderate PM staining with ≥50% circumferential PM coverage in ≥10% of DCIS cells. **(E)** Score 3+: Strong PM staining with ≥50% circumferential PM coverage in ≥10% of DCIS cells. Arrowheads highlight incomplete **(B-C)** versus complete **(D-E)** membrane rims. Scale bars, 100 µm; identical acquisition and display settings. **(F)** Overall PM-TRPV4 score distribution across all 225 scored DCIS image fields from three independent pathologists (675 total ratings). Bars show the proportion of ratings in each score category. **(G)** PM-TRPV4 score distributions stratified by histologic grade (LG, low grade; IMG, intermediate grade; HG, high grade). Bars show the proportion of ratings in each score category within each grade stratum. Higher scores (2+ and 3+) were more frequent in intermediate- and high-grade DCIS. **(H)** PM-TRPV4 positivity by histologic grade at the image level (fields: LG, n=35; IMG, n=139; HG, n=51). Values are proportions of PM-positive fields; red circles indicate mean values and error bars indicate 95% Clopper-Pearson CIs. Trend tests used Cochran-Armitage (z = 4.03, p = 5.6 × 10^-5) and Spearman (ρ = 0.204, p = 0.002).

Score distributions are shown in **Fig. 4F** (overall) and **Fig. 4G** (stratified by grade). Interobserver agreement on the five-level ordinal scale was almost perfect (linearly weighted Fleiss’ κ = 0.823, 95% CI 0.777–0.863, p < 0.001; unweighted Fleiss’ κ = 0.728, 95% CI 0.639–0.818) (**Supplementary Fig. S3**; see **Supplementary Data S2** for indexed scores for all 225 ROIs). Observed agreement was 80.7% (3/3 concordance in 160/225 fields; 2/3 majority in 65/225). Despite mixed subcellular staining patterns, breast pathologists reproducibly identified the plasma-membrane component using the prespecified rubric.

As established in our prior mechanistic work (27), crowding-induced mechanical stress in confined ducts drives TRPV4 relocalization to the plasma membrane; accordingly, PM-TRPV4 serves as a functional readout of pro-invasive mechanotransduction pathway engagement rather than bulk TRPV4 abundance, consistent with our prior finding that total TRPV4 protein level does not predict invasion risk (27). **Fig. 4H** reports image-level PM-TRPV4-positive rates (scored fields, not patient counts): 28.6% (95% CI 16.3-45.1%) in low-grade (n = 35), 67.6% (59.5-74.8%) in intermediate-grade (n = 139), and 74.5% (61.1–84.5%) in high-grade DCIS (n = 51), with a significant increasing trend across grades (Cochran–Armitage z = 4.03, p = 5.6 × 10⁻⁵; Spearman ρ = 0.204, p = 0.002). We therefore examined whether PM-TRPV4 positivity is associated with invasive progression.

### PM-TRPV4 positivity is independently associated with invasive progression-free survival

In the cohort of 44 patients (24 progression events; median follow-up 7.6 years, range 0.4–14.9 years), Kaplan–Meier curves showed visible separation by PM-TRPV4 status (**Fig. 5A**, **Table 2**). PM-TRPV4-positive cases had inferior invasive progression-free survival compared with PM-TRPV4-negative cases (log-rank p = 0.0396). By contrast, conventional histologic grade did not significantly stratify outcome in this dataset (**Fig. 5B**, **Table 2**), with overlapping curves for high-versus lower-grade disease (log-rank p = 0.7199).

**Figure 5.**
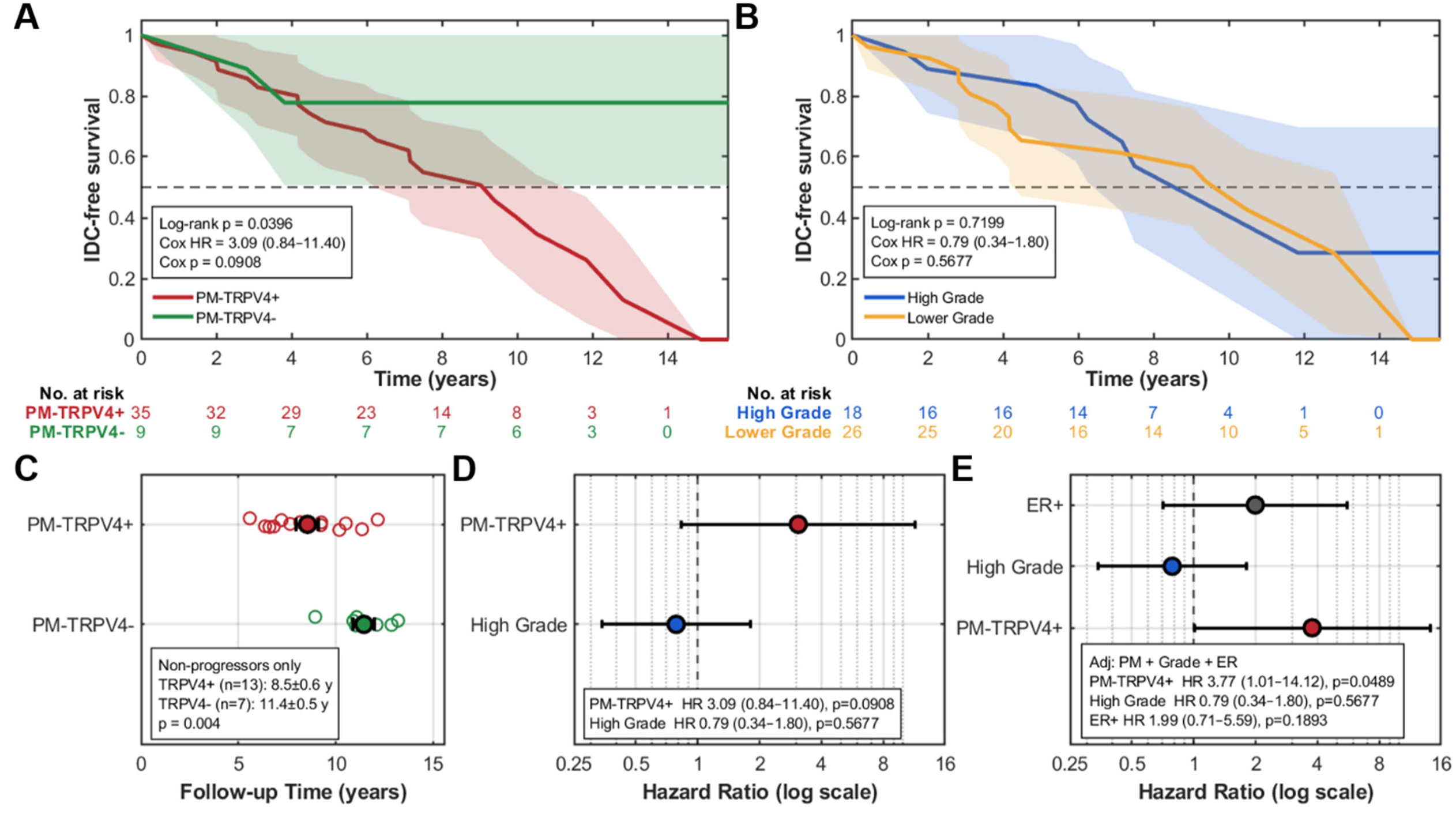
Time to invasive progression by PM-TRPV4 status and histologic grade. Cohort: 44 patients; 24 progression events (54.5%); median follow-up 7.6 y (range 0.4–14.9). **(A)** Kaplan–Meier curves for time to invasive progressionstratified by PM-TRPV4 status in 44 patients (24 events; median follow-up 7.6 y, range 0.4–14.9). PM-TRPV4-positive (red; n=35) had worse survival than PM-TRPV4-negative (green; n=9). Shaded bands are 95% CIs (Greenwood). Dashed line marks 50% survival. Numbers at risk (every 2 y): PM-TRPV4+ 35, 32, 29, 23, 14, 8, 3, 1; PM-TRPV4− 9, 9, 7, 7, 7, 6, 3, 0. Log-rank p=0.0396. Univariable Cox HR=3.09 (95% CI 0.84–11.40), Wald p=0.0908. **(B)** Kaplan–Meier curves by histologic grade (blue=High, n=18; orange=Lower, n=26) with the same conventions. Numbers at risk (every 2 y): High 18, 16, 16, 14, 7, 4, 1, 0; Lower 26, 25, 20, 16, 14, 10, 5, 1. Log-rank p=0.7199. Univariable Cox HR=0.79 (95% CI 0.34–1.80), Wald p=0.5677 (High vs Lower). **(C)** Non-progressors (with ≥5 years of invasive progression-free surveillance): observed surveillance duration by PM-TRPV4 status. PM-TRPV4+: 8.5 ± 0.6 y (n=13); PM-TRPV4-: 11.4 ± 0.5 y (n=7); two-sided Welch t-test p=0.004. **(D)** Univariable Cox forest plot on a log scale (vertical dashed line at reference HR=1). PM-TRPV4+: HR=3.09 (95% CI 0.84–11.40), Wald p=0.0908. High Grade: HR=0.79 (95% CI 0.34–1.80), Wald p=0.5677. **(E)** Multivariable Cox forest plot adjusted for PM-TRPV4, grade, and ER status (log scale; reference HR=1). ER+: adjusted HR=1.99 (95% CI 0.71–5.59), Wald p=0.1893. High Grade: adjusted HR=0.79 (95% CI 0.34–1.80), Wald p=0.5677. PM-TRPV4+: adjusted HR=3.77 (95% CI 1.01–14.12), Wald p=0.0489. PM-TRPV4 positivity remained independently associated with shorter time to invasive progression after adjustment for grade and ER status. PM-TRPV4⁻, lower grade, ER⁻. All p-values are two-sided.

**Table 2.** Cox models for time to invasive progression (N=44; 24 events) Time to invasive progression was analyzed with Cox proportional-hazards regression. For each predictor, we report (i) univariable estimates and their two-sided Wald p-values (Univ p), and (ii) multivariable estimates (Adjusted HR) from a model including PM-TRPV4 status, histologic grade, and ER status; the associated two-sided Wald p-values are shown as Adj p. Hazard ratios (HRs) and 95% confidence intervals (CIs) are reported for each model. Time-to-event was measured from diagnosis to invasive progression or censoring at last follow-up. Ties were handled using the Efron method. All tests were two-sided with α=0.05. (ER was included as an adjustment covariate and was not prespecified for separate univariable testing.)

| **Variable (reference)** | **Univariable HR**<br>**(95% CI)** | **Univ. p** | **Adjusted HR**<br>**(95% CI)** | **Adj. p** |
| --- | --- | --- | --- | --- |
| PM-TRPV4+ (vs -) | 3.09 (0.84–11.40) | 0.0908 | **3.77 (1.01–14.12)** | **0.0489** |
| High grade (vs lower) | 0.79 (0.34–1.80) | 0.5677 | 0.79 (0.34–1.80) | 0.5677 |
| ER+ (vs ER-) | — | — | 1.99 (0.71–5.59) | 0.1893 |

Within the non-progressor subset (invasive progression-free surveillance for ≥5 years), observed surveillance duration was shorter in PM-TRPV4+ than PM-TRPV4− cases (8.5 ± 0.6 vs 11.4 ± 0.5 years; two-sided Welch t-test p = 0.004; **Fig. 5C**). This imbalance would bias against detecting a survival difference, not toward creating one, suggesting the observed association is conservative.

Effect sizes from Cox models are summarized in the forest plots. In univariable Cox (**Fig. 5D**), PM-TRPV4+ was associated with higher hazard of progression (HR = 3.09; 95% CI 0.84–11.40; Wald p = 0.0908), whereas histologic grade was not significant (high vs lower: HR = 0.79; 95% CI 0.34–1.80; Wald p = 0.5677). In a multivariable Cox model including PM-TRPV4, histologic grade, and ER status (**Fig. 5E**, **Table 2**), PM-TRPV4 remained independently associated with shorter invasive progression-free survival (adjusted HR = 3.77; 95% CI 1.01–14.12; Wald p = 0.0489). Neither high grade (adjusted HR = 0.79; 95% CI 0.34–1.80; Wald p = 0.5677) nor ER positivity (adjusted HR = 1.99; 95% CI 0.71–5.59; Wald p = 0.1893) was significant after adjustment. PM-TRPV4 thus added prognostic information that grade and ER status alone did not provide in this cohort.

Proportional-hazards assumptions were assessed using Schoenfeld residuals and satisfied for PM-TRPV4 (p = 0.67) and histologic grade (p = 0.39). Although non-proportionality was detected for ER (p = 0.0001), ER was not prognostic (p = 0.19), so this does not affect interpretation of the primary PM-TRPV4 findings.

Exploratory analyses stratified by histologic grade (**Supplementary Fig S4; Supplementary Table S4**) suggested a stronger association in lower-grade DCIS, whereas estimates in high-grade disease were imprecise because only one PM-TRPV4-negative case was available. In lower-grade disease, PM-TRPV4+ cases had shorter invasive progression-free survival (**Supplementary Fig S4,C**; log-rank p = 0.0703; HR = 3.24; 95% CI 0.85–12.41; Wald p = 0.0859) and a higher progression proportion (14/18 [77.8%] vs 2/8 [25.0%]; Fig. S2D). In high-grade disease, no significant difference was detected (**Supplementary Fig S4B,C**; log-rank p = 0.2280; HR = 1.85; 95% CI 0.11–32.06; Wald p = 0.6723). These subgroup findings are consistent with the primary analysis and support PM-TRPV4 as most clinically informative in lower-grade DCIS, where risk stratification is most challenging.

Univariable analysis (**Supplementary Table S5**) showed that PM-TRPV4 positivity was enriched among progressors (22/24 vs 13/20) with higher odds of progression (OR = 5.92; 95% CI 1.07–32.87; two-sided p = 0.057); this did not reach the α = 0.05 threshold but was directionally consistent with the time-to-event findings. In contrast, histologic grade (OR = 0.50; p = 0.36), ER status (OR = 1.78; p = 0.68), and comorbid factors including cardiovascular disease, metabolic disorder, and smoking history were not associated with progression.

### Exploratory sensitivity analyses of treatment selection and event laterality

To evaluate whether the PM-TRPV4 association might be confounded by treatment selection, we compared PM-TRPV4 status with surgical management (intensive treatment defined as lumpectomy + radiation or mastectomy). PM-TRPV4 was not associated with receipt of intensive therapy versus lumpectomy alone (OR = 2.33; 95% CI 0.45–12.06; two-sided Fisher’s p = 0.367; **Supplementary Table S6**). Surgical intensity also was not associated with progression (OR = 0.50; 95% CI 0.11–2.33; two-sided Fisher’s p = 0.467; **Supplementary Table S7**). Among progressed cases, ipsilateral and contralateral events occurred with similar frequency (13 vs 10; **Supplementary Table S8**). Laterality showed no association with PM-TRPV4 status (two-sided Fisher’s exact p = 0.482; **Supplementary Table S9**). Together, these exploratory analyses did not identify evidence that the PM-TRPV4 signal was explained by treatment choice or event laterality, although the sample size was too small to support treatment-management inferences.

## Discussion

In this single-institution retrospective case-control study of 44 patients with pure DCIS (24 progressors; median follow-up 7.6 years), PM-TRPV4 was associated with shorter time to invasive progression and remained independently prognostic after adjustment for histologic grade and ER status.

TRPV4 distributes across multiple subcellular compartments, including plasma membrane, cytoplasm, and nucleus. Stress-induced PM relocalization directly marks pro-invasive mechanotransduction pathway activation: crowding-induced TRPV4 inactivation triggers compensatory PM accumulation of inactive TRPV4, positioning it for reactivation by future mechanical cues (27). Cytoplasmic and nuclear pools serve as the intracellular reservoir from which this relocalization is drawn (27). In addition, nuclear TRPV4 is biologically regulated through transcriptional and trafficking mechanisms, as reinforced by recent reports (32, 33) and consistent with the bidirectional nuclear-PM trafficking we observe (27). Among these compartments, the PM-relocalized pool is uniquely suited as a clinical readout because it directly indicates mechanosensing pathway engagement. The assay is therefore a functional localization biomarker grounded in the biology of ductal crowding, rather than a measure of total TRPV4 protein abundance or generic membrane expression, which does not correlate with pathologic aggressiveness (27).

The scored PM pattern is supported by orthogonal antibody validation at two levels. At the time of study initiation, no independently KD/KO-validated anti-TRPV4 antibody was available for IHC; Abcam ab39260 was therefore selected based on best performance among antibodies tested via knockdown in our mechanistic work (27). Such a KD/KO-validated antibody has since become available (MilliporeSigma ZRB3155), and we used it to confirm staining specificity on matched material. First, on matched DCIS IHC sections, the orthogonal validation antibody, MilliporeSigma ZRB3155, showed closely concordant compartmental distributions with Abcam ab39260 at the single-cell level, with only 1.8-2.6% deviation across compartment-level metrics. Second, in mechanosensing MCF10DCIS.com cell-line models, both antibodies recapitulated the crowding-induced PM relocalization, shifting from nuclear-dominant patterns under normal-density conditions to strongly PM-dominant patterns under overcrowding, with less than 1% deviation between antibodies under overcrowded conditions. This crowding-induced PM relocalization was absent in non-mechanosensing MCF10A cells (27), confirming specificity of the PM-TRPV4 signal to the mechanosensing context. Together, these validations establish that the scored PM pattern reflects a reproducible mechanosensing-linked signal rather than reagent-specific staining behavior.

These orthogonal validations support the feasibility of PM-TRPV4 scoring, as evidenced by our results. Three breast pathologists scoring 225 image fields achieved almost perfect agreement on the five-level ordinal scale (linearly weighted Fleiss’ κ = 0.823; 95% CI 0.777-0.863), with even higher agreement after dichotomization (unweighted Fleiss’ κ = 0.851). This reproducibility is notable because PM scoring required active discrimination of a membrane rim from mixed subcellular signal, a contextually demanding call made reliably across three independent raters.

These orthogonal validations and high scoring concordance reinforce the clinical findings. Kaplan–Meier curves separated by PM-TRPV4 status (log-rank p = 0.0396), and PM-TRPV4 positivity remained independently linked to shorter time to invasive progression in multivariable Cox regression (adjusted HR = 3.77; 95% CI 1.01–14.12; p = 0.0489). By contrast, conventional histologic grade was not significantly prognostic in this cohort (adjusted HR = 0.79; 95% CI 0.34–1.80; p = 0.5677). In case-control analysis, PM-TRPV4 positivity showed higher odds of progression (OR = 5.92; 95% CI 1.07-32.87; two-sided p = 0.057), directionally consistent with the survival findings but not achieving the α = 0.05 threshold. The strongest signal appeared in lower-grade disease (**Supplementary Fig. S4**; **Supplementary Table S4**): among intermediate and low-grade cases, OR = 10.50 (95% CI 1.54–71.76; p = 0.016) and HR = 3.24 (95% CI 0.85–12.41; p = 0.0859), whereas the high-grade stratum was underpowered due to very few PM-negative cases. Exploratory sensitivity analyses did not identify evidence that the PM-TRPV4 signal was explained by treatment selection, surgical intensity, or event laterality (**Supplementary Tables S5-S9**), although these analyses were limited by small sample size.

This study was designed to ask whether PM-TRPV4 scoring is feasible on routine pathology material and whether a detectable association with invasive progression exists sufficient to justify prospective validation. On both counts the answer is yes. The single-institution design and modest sample size are appropriate for this stage; establishing that the signal exists and the assay is scorable is the necessary precondition for a larger study, not a substitute for one. The non-progressor requirement of ≥5 years invasive progression-free surveillance is conservative by design; the shorter observed surveillance in PM-TRPV4+ non-progressors would blur rather than inflate the survival difference, suggesting the true effect could be stronger with ≥10-year follow-up. Case-control sampling was chosen deliberately to enrich for events; time-to-event measures and ORs, which remain valid under this design, were prespecified accordingly. Analyses were deliberately limited and prespecified to reduce overfitting and overinterpretation, interobserver agreement was high, and findings were directionally consistent across all sensitivity analyses. The study does not claim definitive clinical validation; rather, it establishes proof-of-concept and defines the agenda for prospective multi-institutional evaluation.

Clinically, PM-TRPV4 may complement histologic grading, particularly in lower-grade DCIS where morphology alone may be insufficient for risk stratification. If validated, PM-TRPV4 could help identify biologically active lesions that merit closer monitoring in future studies of risk-stratified management. Immediate next steps include multi-institutional cohorts with prospective follow-up, standardized staining, scoring anchors, and longer-term surveillance; head-to-head comparison with existing clinicopathologic risk models and other established prognostic approaches (34, 35); direct comparison of PM-TRPV4 scoring with established membrane-based IHC scoring frameworks such as HER2 in DCIS (36–39); and integration as an exploratory endpoint in active-surveillance trials. Compatibility with standard FFPE IHC workflows supports the feasibility of broader multi-site implementation with appropriate training and scoring anchors. Our findings provide proof-of-concept that a pathology-readable IHC readout of TRPV4 relocalization, grounded in the mechanosensing biology of ductal crowding, may provide prognostic information beyond grade and ER in DCIS and warrants multi-institutional validation.

## Data Availability

De-identified clinical variables are available from the corresponding author upon reasonable request, subject to institutional data-sharing policies and IRB approval. Anonymized scoring results supporting the findings of this study are provided as Supplementary Data. All analysis scripts are publicly available at the ichung-lab GitHub repository (ichung-lab/Longitudinal, /code).

## Abbreviations

DCIS: ductal carcinoma in situ
IDC: invasive ductal carcinoma
PM: plasma membrane
PM-TRPV4: plasma-membrane TRPV4
TRPV4: transient receptor potential vanilloid 4
ER: estrogen receptor
PR: progesterone receptor
FFPE: formalin-fixed paraffin-embedded
IHC: immunohistochemistry
ROI: region of interest
HG: high grade
HR: hazard ratio
OR: odds ratio
CI: confidence interval
SD: standard deviation
SE: standard error
KM: Kaplan–Meier

## References

1. Allred DC. Ductal carcinoma in situ: terminology, classification, and natural history. J Natl Cancer Inst Monogr. 2010;2010(41):134–8.

2. Bocker W. Preneoplasia of the breast. Verh Dtsch Ges Pathol. 1997;81:502–13.

3. Gudjonsson T, Adriance MC, Sternlicht MD, Petersen OW, Bissell MJ. Myoepithelial cells: Their origin and function in breast morphogenesis and neoplasia. J Mammary Gland Biol. 2005;10(3):261–72.

4. Siegel RL, Kratzer TB, Giaquinto AN, Sung H, Jemal A. Cancer statistics, 2025. CA Cancer J Clin. 2025;75(1):10–45.

5. van Seijen M, Lips EH, Thompson AM, Nik-Zainal S, Futreal A, Hwang ES, et al. Ductal carcinoma in situ: to treat or not to treat, that is the question. Br J Cancer. 2019;121(4):285–92.

6. Kumar AS, Bhatia V, Henderson IC. Overdiagnosis and overtreatment of breast cancer: rates of ductal carcinoma in situ: a US perspective. Breast Cancer Res. 2005;7(6):271–5.

7. Baxter NN, Virnig BA, Durham SB, Tuttle TM. Trends in the treatment of ductal carcinoma in situ of the breast. J Natl Cancer Inst. 2004;96(6):443–8.

8. Srivastava S, Koay EJ, Borowsky AD, De Marzo AM, Ghosh S, Wagner PD, et al. Cancer overdiagnosis: a biological challenge and clinical dilemma. Nat Rev Cancer. 2019;19(6):349–58.

9. Ryser MD, Horton JK, Hwang ES. How Low Can We Go-and Should We? Risk Reduction for Minimal-Volume DCIS. Ann Surg Oncol. 2018;25(2):354–5.

10. Ryser MD, Weaver DL, Zhao F, Worni M, Grimm LJ, Gulati R, et al. Cancer Outcomes in DCIS Patients Without Locoregional Treatment. J Natl Cancer Inst. 2019;111(9):952–60.

11. Hanna WM, Parra-Herran C, Lu FI, Slodkowska E, Rakovitch E, Nofech-Mozes S. Ductal carcinoma in situ of the breast: an update for the pathologist in the era of individualized risk assessment and tailored therapies. Mod Pathol. 2019;32(7):896–915.

12. Worni M, Akushevich I, Greenup R, Sarma D, Ryser MD, Myers ER, et al. Trends in Treatment Patterns and Outcomes for Ductal Carcinoma In Situ. J Natl Cancer Inst. 2015;107(12):djv263.

13. Maxwell AJ, Clements K, Hilton B, Dodwell DJ, Evans A, Kearins O, et al. Risk factors for the development of invasive cancer in unresected ductal carcinoma in situ. Eur J Surg Oncol. 2018;44(4):429–35.

14. Collins LC, Tamimi RM, Baer HJ, Connolly JL, Colditz GA, Schnitt SJ. Outcome of patients with ductal carcinoma in situ untreated after diagnostic biopsy: results from the Nurses’ Health Study. Cancer. 2005;103(9):1778–84.

15. Kerlikowske K, Molinaro A, Cha I, Ljung BM, Ernster VL, Stewart K, et al. Characteristics associated with recurrence among women with ductal carcinoma in situ treated by lumpectomy. J Natl Cancer Inst. 2003;95(22):1692–702.

16. Rakovitch E, Nofech-Mozes S, Hanna W, Baehner FL, Saskin R, Butler SM, et al. A population-based validation study of the DCIS Score predicting recurrence risk in individuals treated by breast-conserving surgery alone. Breast Cancer Res Treat. 2015;152(2):389–98.

17. Erbas B, Provenzano E, Armes J, Gertig D. The natural history of ductal carcinoma in situ of the breast: a review. Breast Cancer Res Treat. 2006;97(2):135–44.

18. Sanders ME, Schuyler PA, Dupont WD, Page DL. The natural history of low-grade ductal carcinoma in situ of the breast in women treated by biopsy only revealed over 30 years of long-term follow-up. Cancer. 2005;103(12):2481–4.

19. Hulahan TS, Spruill L, Wallace EN, Park Y, West RB, Marks JR, et al. Extracellular Microenvironment Alterations in Ductal Carcinoma In Situ and Invasive Breast Cancer Pathologies by Multiplexed Spatial Proteomics. Int J Mol Sci. 2024;25(12).

20. Ma A, Xiang L, Yuan J, Wang Q, Zhao L, Yan H. Spatial Transcriptomics Decodes Breast Cancer Microenvironment Heterogeneity: From Multidimensional Dynamic Profiling to Precision Therapy Blueprint Construction. Biomolecules. 2025;15(8).

21. Strand SH, Houlahan KE, Branch V, King LM, Lynch T, Rivero-Guitiérrez B, et al. Analysis of ductal carcinoma in situ by self-reported race reveals molecular differences related to outcome. Breast Cancer Res. 2024;26(1):127.

22. Hwang ES, Hyslop T, Lynch T, Frank E, Pinto D, Basila D, et al. The COMET (Comparison of Operative versus Monitoring and Endocrine Therapy) trial: a phase III randomised controlled clinical trial for low-risk ductal carcinoma in situ (DCIS). BMJ Open. 2019;9(3):e026797.

23. Elshof LE, Tryfonidis K, Slaets L, van Leeuwen-Stok AE, Skinner VP, Dif N, et al. Feasibility of a prospective, randomised, open-label, international multicentre, phase III, non-inferiority trial to assess the safety of active surveillance for low risk ductal carcinoma in situ - The LORD study. Eur J Cancer. 2015;51(12):1497–510.

24. Francis A, Thomas J, Fallowfield L, Wallis M, Bartlett JM, Brookes C, et al. Addressing overtreatment of screen detected DCIS; the LORIS trial. Eur J Cancer. 2015;51(16):2296–303.

25. Partridge AH, Hyslop T, Rosenberg SM, Bennett AV, Drier S, Jonsson M, et al. Patient-Reported Outcomes for Low-Risk Ductal Carcinoma In Situ: A Secondary Analysis of the COMET Randomized Clinical Trial. JAMA Oncol. 2025;11(3):300–9.

26. Shah C, Bremer T, Cox C, Whitworth P, Patel R, Patel A, et al. The Clinical Utility of DCISionRT(®) on Radiation Therapy Decision Making in Patients with Ductal Carcinoma In Situ Following Breast-Conserving Surgery. Ann Surg Oncol. 2021;28(11):5974–84.

27. Bu X, Ashby N, Vitali T, Lee S, Gottumukkala A, Yun K, et al. Cell crowding activates pro-invasive mechanotransduction pathway in high-grade DCIS via TRPV4 inhibition and cell volume reduction. eLife. 2025:13:RP100490.

28. Rosenbaum T, Benitez-Angeles M, Sanchez-Hernandez R, Morales-Lazaro SL, Hiriart M, Morales-Buenrostro LE, et al. TRPV4: A Physio and Pathophysiologically Significant Ion Channel. Int J Mol Sci. 2020;21(11).

29. White JP, Cibelli M, Urban L, Nilius B, McGeown JG, Nagy I. TRPV4: Molecular Conductor of a Diverse Orchestra. Physiol Rev. 2016;96(3):911–73.

30. Hua R, Jiang JX. How cell crowding causes cancer cells to spread. eLife. 2025;14:e106768.

31. Ashby N, Rubin M, Hawley RG, Chung I. Log-linear scaling of TRPV4-KCNN4 transcripts tunes ROCK-dependent mechanotransduction in a DCIS progression model. bioRxiv. 2026:2026.02.19.706850.

32. Méndez-Gómez S, Espadas-Alvarez H, Ramírez-Rodríguez I, Domínguez-Malfavón L, García-Villegas R. The amino-terminal domain of TRPV4 channel is involved in its trafficking to the nucleus. Biochem Bioph Res Co. 2022;592:13–7.

33. An D, Qi X, Li K, Xu W, Wang Y, Chen X, et al. Blockage of TRPV4 Downregulates the Nuclear Factor-Kappa B Signaling Pathway to Inhibit Inflammatory Responses and Neuronal Death in Mice with Pilocarpine-Induced Status Epilepticus. Cell Mol Neurobiol. 2023;43(3):1283–300.

34. Silverstein MJ. The University of Southern California/Van Nuys prognostic index for ductal carcinoma in situ of the breast. The American Journal of Surgery. 2003;186(4):337–43.

35. Weinmann S, Leo MC, Francisco M, Jenkins CL, Barry T, Leesman G, et al. Validation of a Ductal Carcinoma In Situ Biomarker Profile for Risk of Recurrence after Breast-Conserving Surgery with and without Radiotherapy. Clin Cancer Res. 2020;26(15):4054–63.

36. Miligy IM, Toss MS, Gorringe KL, Lee AHS, Ellis IO, Green AR, et al. The clinical and biological significance of HER2 over-expression in breast ductal carcinoma in situ: a large study from a single institution. British Journal of Cancer. 2019;120(11):1075–82.

37. Van Bockstal MR, Wesseling J, Lips EH, Smidt M, Galant C, van Deurzen CHM. Systematic assessment of HER2 status in ductal carcinoma in situ of the breast: a perspective on the potential clinical relevance. Breast Cancer Research. 2024;26(1):125.

38. Thorat MA, Levey PM, Jones JL, Pinder SE, Bundred NJ, Fentiman IS, et al. Prognostic and Predictive Value of HER2 Expression in Ductal Carcinoma: Results from the UK/ANZ DCIS Randomized Trial. Clinical Cancer Research. 2021;27(19):5317–24.

39. Wolff AC, Somerfield MR, Dowsett M, Hammond MEH, Hayes DF, McShane LM, et al. Human Epidermal Growth Factor Receptor 2 Testing in Breast Cancer. Arch Pathol Lab Med. 2023;147(9):993–1000.

