## Supplementary Figures and Tables for "Analytical validation and proof-of-concept prognostic evaluation of plasma-membrane TRPV4 localization in ductal carcinoma in situ: a retrospective case-control study"

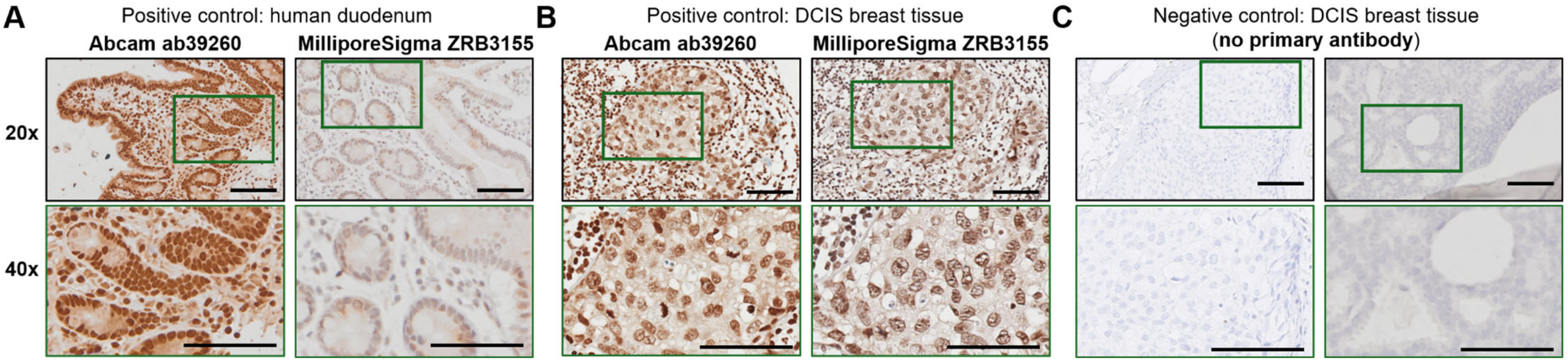


**Supplementary Figure S1. Comparative TRPV4 immunohistochemistry with Abcam ab39260 and MilliporeSigma ZRB3155 antibodies in control tissues.**

**(A)** Human duodenum positive-control tissue stained with primary study antibody (Abcam ab39260) or the orthogonal validation antibody MilliporeSigma ZRB3155.

**(B)** Positive control DCIS breast tissue stained with Abcam ab39260 or MilliporeSigma ZRB3155. For each antibody, 20× views and corresponding 40× magnified regions are shown.

**(C)** Negative-control DCIS breast tissue processed in parallel without primary antibody. Green boxes indicate the areas shown at higher magnification. Both antibodies demonstrate positive staining in control tissues, whereas omission of the primary antibody yields minimal background, supporting assay specificity under the applied staining conditions.

Scale bars, 100 μm.

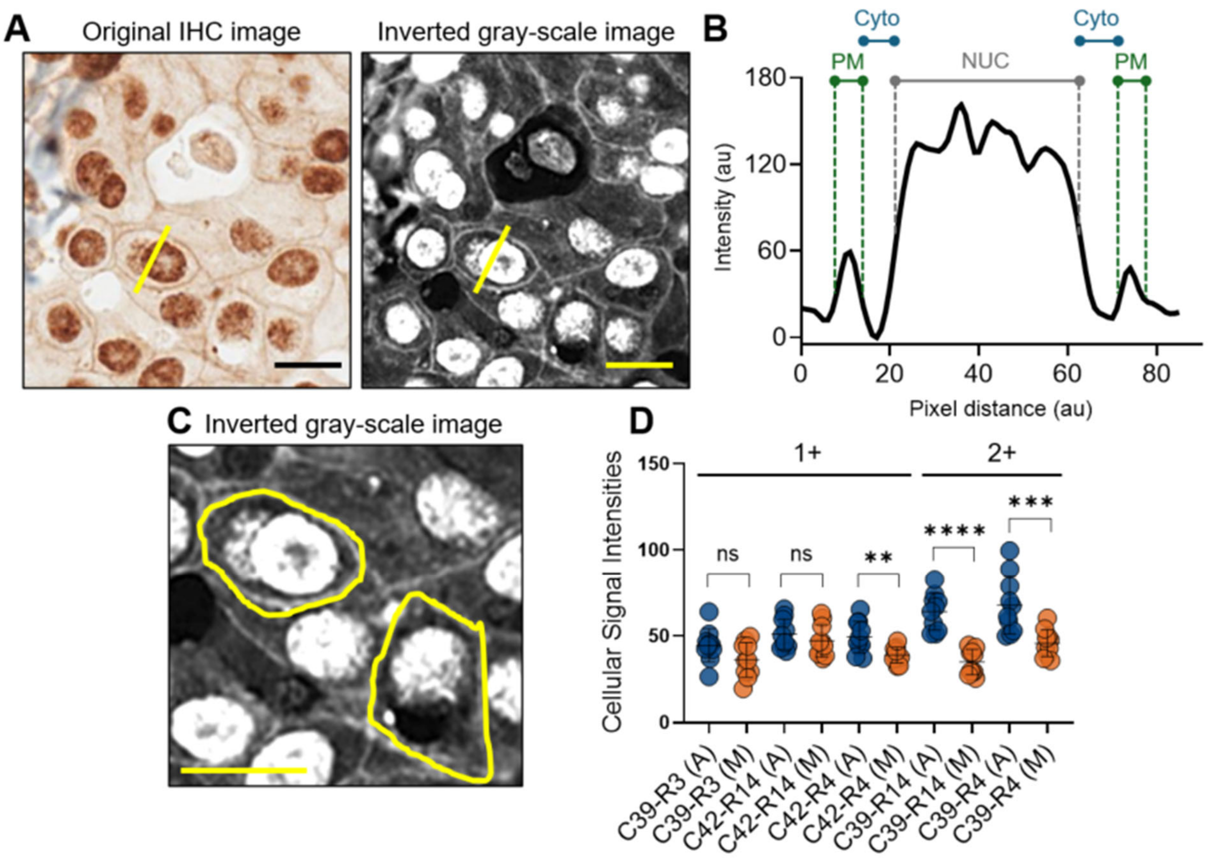


**Supplementary Figure S2. Single-cell line analysis for subcellular TRPV4 compartment quantification and cell staining intensity measurements.**

**(A)** Line analysis method: Representative TRPV4 IHC image (Abcam ab39260) of a scored DCIS ROI (left) and the inverted 8-bit image (right). A straight yellow line is drawn across a single epithelial cell spanning from one plasma-membrane (PM) edge to the opposite PM edge, passing through the cytoplasm (Cyto) and nucleus (NUC). The line is positioned to cross the cell to ensure all three compartments are sampled.

**(B)** Intensity profile. Background-subtracted pixel intensity along the line defined in **(A)**, plotted as a function of position. Peaks at the left and right margins correspond to plasma membrane (PM; green) signal; the central peak corresponds to nuclear (NUC; gray) signal; intervening regions reflect cytoplasmic signal (Cyto; teal). Compartment boundaries were defined at the 50% intensity-height threshold separating the three regions.

**(C)** Cell intensity estimation. Individual epithelial cells with clear PM rims were manually segmented to estimate background-subtracted total single-cell intensity for each antibody, enabling direct comparison of absolute staining magnitude independent of compartment fractions.**(D)** Cell-level signal intensities are summarized for 10 cells per ROI across five paired ROIs. Absolute whole-ROI intensity values were evaluated separately after background subtraction and are provided in **Supplementary Table S2**.

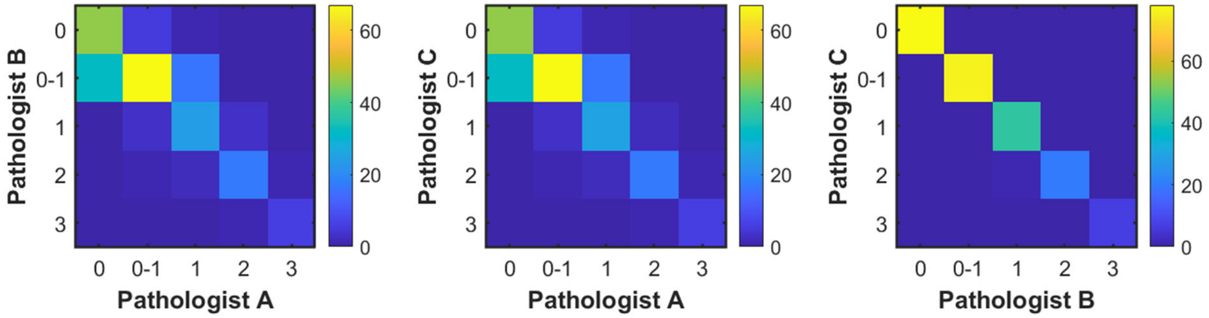


**Supplementary Figure S3. Inter-rater agreement metrics and confusion matrices**

**(A)** Pairwise confusion matrices showing agreement between pathologists for the 5-level PM-TRPV4 scoring (Pathologist A vs B, A vs C, B vs C). Cell colors indicate frequency of rating combinations.

**(B)** Summary of inter-rater reliability metrics. Linear-weighted Fleiss' $\kappa=0.823$ (95% CI 0.777–0.863, $p<0.001$); unweighted Fleiss' $\kappa=0.728$ (95% CI 0.639–0.818, $p<0.001$). Overall observed agreement was 80.7%, with exact 3/3 rater concordance in 71.1% of cases (160/225 fields) and 2/3 majority agreement in 28.9% (65/225 fields). Pairwise weighted Cohen's $\kappa$: A-B 0.731, A-C 0.734, B-C 0.996 (range: 0.731 to 0.996). After dichotomization to PM+/PM-, binary Fleiss' $\kappa=0.851$.

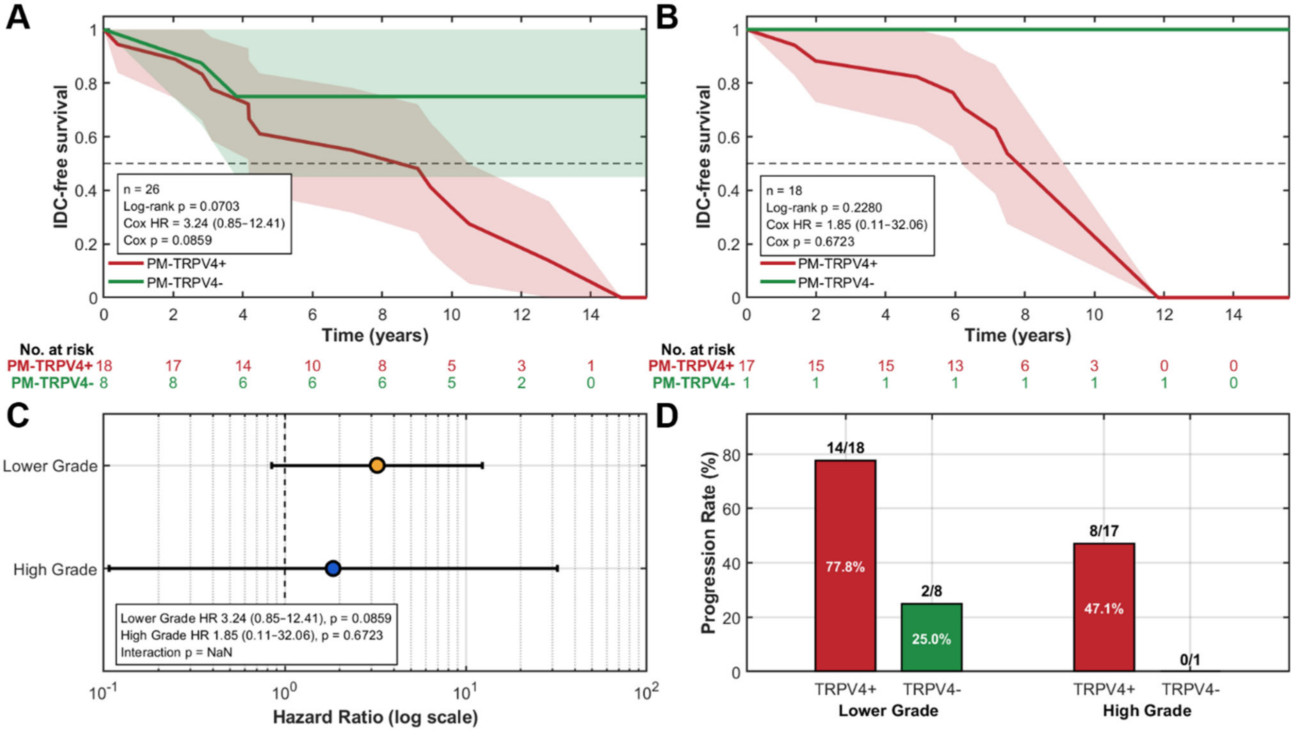


**Supplementary Figure S4. PM-TRPV4 effect within histologic-grade subgroups**

**(A)** Cohort: Same 44 patients from Figure 3, stratified by grade. Kaplan–Meier curves for time to invasive progression among lower-grade DCIS (n=26). PM-TRPV4-positive patients (red) show poorer survival than PM-TRPV4-negative patients (green); shaded bands are 95% CIs (Greenwood), dashed line marks 50% survival, and numbers at risk are shown every 2 years. Log-rank p=0.0703. Univariable Cox: HR 3.24 (95% CI 0.85–12.41), p=0.0859 (PM-TRPV4+ vs PM-TRPV4−).

**(B)** Kaplan–Meier curves for high-grade DCIS (n=18) with the same display conventions. Log-rank p=0.2280. Univariable Cox: HR 1.85 (95% CI 0.11–32.06), p=0.6723.

**(C)** Forest plot summarizing the PM-TRPV4 effect within each grade on a log-scale hazard-ratio axis (reference HR=1). Points are HRs; bars are 95% CIs. A formal PM × Grade interaction could not be reliably estimated due to zero events in the High-grade/PM-TRPV4- group.

**(D)** Invasive progression rates by grade and PM-TRPV4 status with counts over bars: lower grade 14/18 (77.8%) vs 2/8 (25.0%); high grade 8/17 (47.1%) vs 0/1 (0%).

| Case# | Progression (1=Yes,0=No) | High grade at time 0 | PM-TRPV4 | Age | ER | PR | Cardiovascular disease | Smoking history | Metabolic disease | Surgical method (DCIS) | Disease site | Time to invasive progression or follow-up (days) |
|---|---|---|---|---|---|---|---|---|---|---|---|---|
| 1 | 1 | 1 | + | 51-55 | 1 | NA | 1 | 1 | 0 | Mastectomy | Ipsilateral | 2166 |
| 2 | 1 | 0 | + | 66-70 | 1 | NA | 1 | 0 | 1 | Mastectomy | Ipsilateral | 1032 |
| 3 | 1 | 0 | + | 51-55 | 1 | NA | 0 | 0 | 0 | Mastectomy | Contralateral | 3627 |
| 4 | 1 | 0 | + | 56-60 | 0 | NA | 1 | 1 | 1 | Lumpectomy + Radiation | Ipsilateral | 5429 |
| 5 | 1 | 1 | + | 56-60 | NA | NA | 1 | 1 | 0 | Mastectomy | Contralateral | 1786 |
| 6 | 1 | 0 | − | 51-55 | 1 | NA | 0 | 0 | 0 | Lumpectomy + Radiation | Ipsilateral | 1024 |
| 7 | 1 | 0 | + | 56-60 | 0 | 0 | 1 | 1 | 1 | Lumpectomy + Radiation | Contralateral | 3429 |
| 8 | 1 | 0 | + | 61-65 | 1 | NA | 1 | 0 | 0 | Lumpectomy + Radiation | Contralateral | 4675 |
| 9 | 1 | 0 | + | 76-80 | 1 | NA | 1 | 1 | 1 | Lumpectomy + Radiation | Contralateral | 3296 |
| 10 | 1 | 1 | + | 66-70 | 1 | NA | 1 | 0 | 0 | Lumpectomy | NA | 2612 |
| 11 | 1 | 1 | + | 61-65 | 1 | NA | 0 | 1 | 0 | Lumpectomy + Radiation | Ipsilateral | 4321 |
| 12 | 1 | 1 | + | 76-80 | 1 | NA | 1 | 1 | 1 | Lumpectomy | Ipsilateral | 2737 |

| | | | | | | | | | | | | |
|---|---|---|---|---|---|---|---|---|---|---|---|---|
| 13 | 1 | 0 | + | 36-40 | 1 | 1 | 1 | 0 | 0 | Lumpectomy | Contralateral | 3835 |
| 14 | 1 | 0 | + | 76-80 | 1 | NA | 1 | 1 | 0 | Lumpectomy | Ipsilateral | 1133 |
| 15 | 1 | 0 | + | 41-45 | 1 | NA | 0 | 0 | 0 | Lumpectomy + Radiation | Ipsilateral | 2597 |
| 16 | 1 | 1 | + | 61-65 | 0 | NA | 0 | 1 | 0 | Mastectomy | Contralateral | 2283 |
| 17 | 1 | 0 | + | 41-45 | 1 | NA | 0 | 0 | 0 | Lumpectomy + Radiation | Ipsilateral | 1518 |
| 18 | 1 | 0 | + | 76-80 | 1 | NA | 1 | 0 | 0 | Lumpectomy + Radiation | Contralateral | 750 |
| 19 | 1 | 1 | + | 61-65 | 1 | NA | 0 | 0 | 1 | Lumpectomy + Radiation | Ipsilateral | 505 |
| 20 | 1 | 0 | + | 66-70 | 1 | NA | 1 | 0 | 1 | Lumpectomy + Radiation | Contralateral | 1524 |
| 21 | 1 | 0 | + | 61-65 | 1 | NA | 0 | 0 | 0 | NA | Ipsilateral | 1636 |
| 22 | 1 | 1 | + | 66-70 | 1 | NA | 1 | 1 | 0 | Mastectomy | Ipsilateral | 726 |
| 23 | 1 | 0 | − | 61-65 | 1 | 1 | 1 | 0 | 1 | Lumpectomy | Ipsilateral | 1394 |
| 24 | 1 | 0 | + | 61-65 | 1 | NA | 1 | 0 | 1 | Lumpectomy | Contralateral | 146 |
| 25 | 0 | 1 | + | 41-45 | 1 | NA | 0 | 0 | 0 | Mastectomy | NA | 2327 |
| 26 | 0 | 1 | − | 61-65 | 1 | NA | 1 | 1 | 0 | Mastectomy | NA | 4822 |
| 27 | 0 | 0 | − | 61-65 | 1 | NA | 1 | 1 | 1 | Lumpectomy + Radiation | NA | 4698 |

| | | | | | | | | | | | | |
|---|---|---|---|---|---|---|---|---|---|---|---|---|
| 28 | 0 | 0< | − | 66-70 | 1 | NA | 1 | 1 | 0 | Lumpectomy | NA | 4418 |
| 29 | 0 | 0 | + | 66-70 | 1 | NA | 1 | 0 | 0 | Lumpectomy + Radiation | NA | 4440 |
| 30 | 0 | 1 | + | 46-50 | 1 | NA | 1 | 1 | 0 | Lumpectomy | NA | 2634 |
| 31 | 0 | 0 | + | 71-75 | NA | NA | 1 | 0 | 0 | Mastectomy | NA | 4145 |
| 32 | 0 | 0 | − | 76-80 | 1 | NA | 0 | 1 | 0 | Lumpectomy + Radiation | NA | 4030 |
| 33 | 0 | 0 | − | 76-80 | 1 | NA | 0 | 0 | 1 | Lumpectomy + Radiation | NA | 3982 |
| 34 | 0 | 1 | + | 66-70 | 0 | NA | 0 | 0 | 1 | Lumpectomy + Radiation | NA | 3841 |
| 35 | 0 | 0 | + | 71-75 | 1 | NA | 1 | 0 | 0 | Lumpectomy + Radiation | NA | 2798 |
| 36 | 0 | 1 | + | 61-65 | 1 | NA | 1 | 1 | 1 | Lumpectomy + Radiation | NA | 3720 |
| 37 | 0 | 0 | − | 51-55 | 1 | NA | 0 | 1 | 0 | Lumpectomy + Radiation | Contralateral | 4045 |
| 38 | 0 | 1 | + | 61-65 | 0 | NA | 1 | 0 | 0 | Lumpectomy + Radiation | NA | 3380 |
| 39 | 0 | 1 | + | 46-50 | 0 | 0 | 0\ | 0 | 0 | Mastectomy | NA | 3373 |
| 40 | 0 | 0 | − | 71-75 | 1 | 1 | 1 | 0 | 1 | Lumpectomy | Ipsilateral | 3267 |
| 41 | 0 | 1 | + | 51-55 | 1 | NA | 1 | 1 | 0 | Lumpectomy + Radiation | NA | 2986 |
| 42 | 0 | 1 | + | 46-50 | 0 | NA | 0 | 0 | 1 | Lumpectomy + Radiation | NA | 2490 |

| | | | | | | | | | | | | |
|---|---|---|---|---|---|---|---|---|---|---|---|---|
| 43 | 0 | 1 | + | 46-50 | 1 | NA | 0 | 1 | 0 | Mastectomy | NA | 2408 |
| 44 | 0 | 0 | + | 66-70 | 1 | NA | 1 | 0 | 0 | Lumpectomy + Radiation | NA | 2040 |

**Supplementary Table S1. Case-level dataset and clinical covariates**

Case-level listing for the DCIS cohort (N = 44). Codes: Progression (1 = progressed to IDC; 0 = non-progressed); HG (1 = high grade; 0 = lower grade); PM-TRPV4 reported as “+”/“−”; ER/PR coded as 1/0 when available; Surgical method categorized as Lumpectomy, Lumpectomy + Radiation, or Mastectomy; Disease site recorded as Ipsilateral/Contralateral when applicable; Time reported as days to IDC or days to last follow-up for non-progressed cases. NA = not available.

| C39-R14 (A) | | | | | | | | | | | | |
|---|---|---|---|---|---|---|---|---|---|---|---|---|
| | Cell 1 | Cell 2 | Cell 3 | Cell 4 | Cell 5 | Cell 6 | Cell 7 | Cell 8 | Cell 9 | Cell 10 | Average | Std Dev |
| PM | 19.14% | 20.69% | 22.13% | 24.18% | 20.65% | 21.55% | 22.93% | 25.91% | 22.89% | 21.99% | 22.21% | 1.92% |
| Nuc | 53.81% | 53.86% | 55.22% | 47.10% | 49.74% | 58.64% | 51.99% | 45.27% | 51.50% | 57.37% | 52.45% | 4.25% |
| Cytosolic | 27.05% | 25.45% | 22.65% | 28.72% | 29.61% | 19.81% | 25.08% | 28.82% | 25.61% | 20.64% | 25.34% | 3.41% |
| Single Cell Signal Intensity | 70.60 | 57.40 | 51.20 | 74.01 | 69.87 | 66.80 | 51.18 | 82.68 | 64.02 | 53.06 | 64.08 | 10.68 |
| Whole ROI Signal Intensity | 52.37 | | | | | | | | | | | |
| PM:Nuc | 0.36 | 0.38 | 0.40 | 0.51 | 0.42 | 0.37 | 0.44 | 0.57 | 0.44 | 0.38 | 0.43 | 0.07 |

| C39-R14 (M) | | | | | | | | | | | | |
|---|---|---|---|---|---|---|---|---|---|---|---|---|
| | Cell 1 | Cell 2 | Cell 3 | Cell 4 | Cell 5 | Cell 6 | Cell 7 | Cell 8 | Cell 9 | Cell 10 | Average | Std Dev |
| PM | 27.85% | 26.50% | 28.91% | 21.13% | 24.47% | 24.60% | 21.67% | 25.44% | 24.65% | 23.28% | 24.85% | 2.48% |
| Nuc | 50.38% | 54.50% | 52.59% | 57.34% | 50.85% | 52.09% | 49.88% | 54.68% | 50.15% | 53.66% | 52.61% | 2.43% |
| Cytosolic | 21.77% | 19.00% | 18.49% | 21.53% | 24.68% | 23.31% | 28.45% | 21.88% | 25.20% | 23.06% | 22.74% | 2.94% |
| Single Cell Signal Intensity | 25.52 | 29.28 | 29.23 | 29.38 | 39.21 | 40.28 | 44.65 | 43.41 | 40.63 | 27.44 | 34.90 | 7.35 |
| Whole ROI Signal Intensity | 29.31 | | | | | | | | | | | |
| PM:Nuc | 0.55 | 0.49 | 0.55 | 0.37 | 0.48 | 0.47 | 0.43 | 0.47 | 0.49 | 0.43 | 0.47 | 0.05 |

| C39-R4 (A) | | | | | | | | | | | | |
|---|---|---|---|---|---|---|---|---|---|---|---|---|
| | Cell 1 | Cell 2 | Cell 3 | Cell 4 | Cell 5 | Cell 6 | Cell 7 | Cell 8 | Cell 9 | Cell 10 | Average | Std Dev |
| PM | 21.43% | 22.34% | 28.64% | 26.60% | 24.90% | 22.42% | 20.06% | 24.13% | 21.41% | 17.53% | 22.95% | 3.23% |
| Nuc | 52.38% | 48.96% | 51.63% | 53.83% | 50.68% | 52.36% | 57.09% | 52.62% | 48.45% | 69.07% | 53.71% | 5.92% |
| Cytosolic | 26.19% | 28.70% | 19.73% | 19.57% | 24.42% | 25.22% | 22.86% | 23.25% | 30.13% | 13.40% | 23.35% | 4.88% |
| Single Cell Signal Intensity | 99.60 | 58.89 | 49.89 | 67.36 | 50.92 | 78.82 | 70.06 | 52.97 | 88.77 | 61.21 | 67.85 | 16.75 |
| Whole ROI Signal Intensity | 54.00 | | | | | | | | | | | |
| PM:Nuc | 0.41 | 0.46 | 0.55 | 0.49 | 0.49 | 0.43 | 0.35 | 0.46 | 0.44 | 0.25 | 0.43 | 0.08 |

| C39-R4 (M) | | | | | | | | | | | | |
|---|---|---|---|---|---|---|---|---|---|---|---|---|
| | Cell 1 | Cell 2 | Cell 3 | Cell 4 | Cell 5 | Cell 6 | Cell 7 | Cell 8 | Cell 9 | Cell 10 | Average | Std Dev |

| | | | | | | | | | | | | |
|---|---|---|---|---|---|---|---|---|---|---|---|---|
| PM | 26.30% | 17.84% | 16.90% | 27.09% | 16.25% | 17.85% | 23.32% | 21.44% | 14.70% | 21.82% | 20.35% | 4.30% |
| Nuc | 53.06% | 57.54% | 61.49% | 55.31% | 55.98% | 49.51% | 49.76% | 56.81% | 69.00% | 51.11% | 55.96% | 5.93% |
| Cytosolic | 20.65% | 24.62% | 21.61% | 17.60% | 27.76% | 32.64% | 26.92% | 21.74% | 16.30% | 27.07% | 23.69% | 5.04% |
| Single Cell Signal Intensity | 43.25 | 47.81 | 39.07 | 42.79 | 35.66 | 60.25 | 49.48 | 36.91 | 48.78 | 54.03 | 45.80 | 7.79 |
| Whole ROI Signal Intensity | 38.14 | | | | | | | | | | | |
| PM:Nuc | 0.50 | 0.31 | 0.27 | 0.49 | 0.29 | 0.36 | 0.47 | 0.38 | 0.21 | 0.43 | 0.37 | 0.10 |
| C39-R3 (A) | | | | | | | | | | | | |
| | Cell 1 | Cell 2 | Cell 3 | Cell 4 | Cell 5 | Cell 6 | Cell 7 | Cell 8 | Cell 9 | Cell 10 | Average | Std Dev |
| PM | 11.15% | 13.99% | 19.54% | 15.51% | 19.20% | 16.14% | 19.32% | 17.80% | 12.57% | 23.80% | 16.90% | 3.79% |
| Nuc | 63.71% | 66.80% | 57.52% | 66.00% | 50.71% | 69.37% | 65.75% | 70.53% | 62.30% | 61.59% | 63.43% | 5.87% |
| Cytosolic | 25.14% | 19.20% | 22.93% | 18.49% | 30.09% | 14.50% | 14.93% | 11.66% | 25.13% | 14.61% | 19.67% | 5.96% |
| Single Cell Signal Intensity | 51.12 | 46.77 | 36.93 | 45.19 | 63.91 | 26.45 | 42.23 | 42.42 | 45.55 | 44.34 | 44.49 | 9.54 |
| Whole ROI Signal Intensity | 42.76 | | | | | | | | | | | |
| PM:Nuc | 0.18 | 0.21 | 0.34 | 0.24 | 0.38 | 0.23 | 0.29 | 0.25 | 0.20 | 0.39 | 0.27 | 0.08 |
| C39-R3 (M) | | | | | | | | | | | | |
| | Cell 1 | Cell 2 | Cell 3 | Cell 4 | Cell 5 | Cell 6 | Cell 7 | Cell 8 | Cell 9 | Cell 10 | Average | Std Dev |
| PM | 17.74% | 19.18% | 14.73% | 17.74% | 18.87% | 22.15% | 17.16% | 19.04% | 16.96% | 20.15% | 18.37% | 2.01% |
| Nuc | 60.46% | 58.84% | 52.23% | 61.55% | 56.64% | 63.32% | 67.01% | 55.23% | 63.39% | 52.40% | 59.11% | 4.94% |
| Cytosolic | 21.80% | 21.98% | 33.04% | 20.71% | 24.50% | 14.53% | 15.83% | 25.73% | 19.65% | 27.45% | 22.52% | 5.48% |
| Single Cell Signal Intensity | 47.44 | 49.71 | 35.74 | 29.37 | 44.26 | 19.50 | 29.42 | 35.89 | 25.94 | 44.05 | 36.13 | 10.07 |
| Whole ROI Signal Intensity | 30.63 | | | | | | | | | | | |
| PM:Nuc | 0.29 | 0.33 | 0.28 | 0.29 | 0.33 | 0.35 | 0.26 | 0.34 | 0.27 | 0.38 | 0.31 | 0.04 |
| C42-R14 (A) | | | | | | | | | | | | |
| | Cell 1 | Cell 2 | Cell 3 | Cell 4 | Cell 5 | Cell 6 | Cell 7 | Cell 8 | Cell 9 | Cell 10 | Average | Std Dev |
| PM | 26.44% | 29.79% | 23.13% | 31.57% | 24.37% | 25.54% | 27.34% | 22.87% | 25.12% | 29.89% | 26.61% | 2.99% |
| Nuc | 54.42% | 57.35% | 45.23% | 53.97% | 54.21% | 61.71% | 60.38% | 52.43% | 56.60% | 54.39% | 55.07% | 4.55% |

| | | | | | | | | | | | | |
|---|---|---|---|---|---|---|---|---|---|---|---|---|
| Cytosolic | 21.31% | 12.86% | 31.64% | 14.46% | 21.42% | 12.76% | 12.27% | 24.70% | 18.28% | 15.72% | 18.54% | 6.28% |
| Single Cell Signal Intensity | 44.14 | 44.00 | 53.27 | 41.03 | 65.51 | 61.93 | 42.99 | 59.62 | 50.31 | 46.08 | 50.89 | 8.78 |
| Whole ROI Signal Intensity | 49.03 | | | | | | | | | | | |
| PM:Nuc | 0.49 | 0.52 | 0.51 | 0.58 | 0.45 | 0.41 | 0.45 | 0.44 | 0.44 | 0.55 | 0.48 | 0.06 |
| C42-R14 (M) | | | | | | | | | | | | |
| | Cell 1 | Cell 2 | Cell 3 | Cell 4 | Cell 5 | Cell 6 | Cell 7 | Cell 8 | Cell 9 | Cell 10 | Average | Std Dev |
| PM | 24.86% | 25.75% | 22.32% | 24.44% | 21.44% | 23.82% | 23.78% | 22.49% | 25.34% | 25.17% | 23.94% | 1.45% |
| Nuc | 51.12% | 56.73% | 49.19% | 53.62% | 46.44% | 47.18% | 43.36% | 52.19% | 53.65% | 54.53% | 50.80% | 4.18% |
| Cytosolic | 24.02% | 17.52% | 28.49% | 21.94% | 32.12% | 29.00% | 32.85% | 25.31% | 21.02% | 20.30% | 25.26% | 5.21% |
| Single Cell Signal Intensity | 36.84 | 60.27 | 40.16 | 39.64 | 38.57 | 46.54 | 44.28 | 46.5 | 63.11 | 55.5 | 47.14 | 9.37 |
| Whole ROI Signal Intensity | 42.84 | | | | | | | | | | | |
| PM:Nuc | 0.49 | 0.45 | 0.45 | 0.46 | 0.46 | 0.50 | 0.55 | 0.43 | 0.47 | 0.46 | 0.47 | 0.03 |
| C42-R4 (A) | | | | | | | | | | | | |
| | Cell 1 | Cell 2 | Cell 3 | Cell 4 | Cell 5 | Cell 6 | Cell 7 | Cell 8 | Cell 9 | Cell 10 | Average | Std Dev |
| PM | 24.59% | 22.84% | 29.77% | 34.95% | 26.91% | 28.35% | 24.60% | 30.11% | 20.28% | 31.32% | 27.37% | 4.39% |
| Nuc | 55.33% | 45.70% | 46.28% | 54.12% | 54.86% | 43.88% | 55.61% | 59.75% | 52.21% | 46.31% | 51.41% | 5.42% |
| Cytosolic | 20.08% | 31.47% | 23.95% | 10.93% | 18.23% | 27.78% | 19.79% | 10.14% | 27.52% | 22.37% | 21.23% | 6.97% |
| Single Cell Signal Intensity | 43.66 | 44.86 | 37.12 | 65.34 | 58.17 | 45.89 | 49.00 | 37.89 | 57.80 | 53.01 | 49.27 | 9.21 |
| Whole ROI Signal Intensity | 50.43 | | | | | | | | | | | |
| PM:Nuc | 0.44 | 0.50 | 0.64 | 0.65 | 0.49 | 0.65 | 0.44 | 0.50 | 0.39 | 0.68 | 0.54 | 0.10 |
| C42-R4 (M) | | | | | | | | | | | | |
| | Cell 1 | Cell 2 | Cell 3 | Cell 4 | Cell 5 | Cell 6 | Cell 7 | Cell 8 | Cell 9 | Cell 10 | Average | Std Dev |
| PM | 25.87% | 32.26% | 22.16% | 22.25% | 26.36% | 26.71% | 24.29% | 24.86% | 27.57% | 22.10% | 25.44% | 3.12% |
| Nuc | 54.76% | 56.94% | 51.80% | 49.20% | 52.70% | 50.16% | 52.65% | 49.57% | 52.63% | 54.39% | 52.48% | 2.45% |
| Cytosolic | 19.37% | 10.80% | 26.04% | 28.54% | 20.94% | 23.13% | 23.06% | 25.56% | 19.79% | 23.52% | 22.08% | 4.89% |
| Single Cell Signal Intensity | 37.75 | 38.12 | 43.38 | 32.40 | 37.72 | 42.50 | 36.72 | 32.68 | 41.16 | 47.05 | 38.95 | 4.64 |

| | | | | | | | | | | | |
|---|---|---|---|---|---|---|---|---|---|---|---|
| Whole ROI Signal Intensity | 38.61 | | | | | | | | | | |
| PM:Nuc | 0.47 | 0.57 | 0.43 | 0.45 | 0.50 | 0.53 | 0.46 | 0.50 | 0.52 | 0.41 | 0.48 | 0.05 |

**Supplementary Table S2. Single-cell compartment fractions, total cell intensities, and PM:nuclear ratios in matched ROIs stained with Abcam ab39260 and MilliporeSigma ZRB3155.**

For each matched ROI, values are reported for 10 manually segmented epithelial cells, including plasma-membrane (PM), nuclear, and cytoplasmic signal fractions, background-subtracted total single-cell intensity, whole-ROI background-subtracted intensity, and PM:nuclear signal ratio. Means and standard deviations are shown for each ROI-antibody pair. Abcam ab39260 is denoted as A and MilliporeSigma ZRB3155 as M. These data support the orthogonal antibody comparison summarized in **Fig. 2** and **Supplementary Fig. S2**.

| OC DCIS (A) | | | | | | | | | | | | |
|---|---|---|---|---|---|---|---|---|---|---|---|---|
| | Cell 1 | Cell 2 | Cell 3 | Cell 4 | Cell 5 | Cell 6 | Cell 7 | Cell 8 | Cell 9 | Cell 10 | Average | Std Dev |
| PM | 44.29% | 54.21% | 46.32% | 47.01% | 42.67% | 50.49% | 61.19% | 46.68% | 48.56% | 42.39% | 48.38% | 5.74% |
| Nuc | 19.17% | 8.80% | 14.30% | 16.72% | 23.12% | 16.48% | 8.76% | 23.44% | 21.20% | 26.41% | 17.84% | 6.01% |
| Cytosolic | 36.53% | 36.99% | 39.38% | 36.26% | 34.20% | 33.03% | 30.06% | 29.88% | 30.24% | 31.20% | 33.78% | 3.41% |
| PM:Nuc | 2.31 | 6.16 | 3.24 | 2.81 | 1.85 | 3.06 | 6.99 | 1.99 | 2.29 | 1.61 | 3.23 | 1.85 |
| OC DCIS (M) | | | | | | | | | | | | |
| | Cell 1 | Cell 2 | Cell 3 | Cell 4 | Cell 5 | Cell 6 | Cell 7 | Cell 8 | Cell 9 | Cell 10 | Average | Std Dev |
| PM | 51.86% | 47.63% | 54.14% | 64.47% | 41.76% | 43.93% | 60.88% | 52.11% | 50.60% | 37.15% | 50.45% | 8.34% |
| Nuc | 18.13% | 26.35% | 22.71% | 9.08% | 8.71% | 25.32% | 12.87% | 19.43% | 17.70% | 24.02% | 18.43% | 6.45% |
| Cytosolic | 30.01% | 26.02% | 23.15% | 26.45% | 49.53% | 30.74% | 26.25% | 28.46% | 31.70% | 38.82% | 31.11% | 7.76% |
| PM:Nuc | 2.86 | 1.81 | 2.38 | 7.10 | 4.79 | 1.73 | 4.73 | 2.68 | 2.86 | 1.55 | 3.25 | 1.77 |
| ND DCIS (A) | | | | | | | | | | | | |
| | Cell 1 | Cell 2 | Cell 3 | Cell 4 | Cell 5 | Cell 6 | Cell 7 | Cell 8 | Cell 9 | Cell 10 | Average | Std Dev |
| PM | 20.49% | 14.27% | 16.42% | 30.71% | 8.96% | 25.21% | 27.13% | 35.45% | 24.17% | 16.89% | 21.97% | 8.08% |
| Nuc | 55.57% | 63.59% | 63.83% | 38.85% | 68.57% | 40.63% | 37.04% | 47.21% | 48.35% | 21.82% | 48.55% | 14.59% |
| Cytosolic | 23.94% | 22.14% | 19.74% | 30.44% | 22.48% | 34.16% | 35.83% | 17.34% | 27.48% | 61.28% | 29.48% | 12.71% |
| PM:Nuc | 0.37 | 0.22 | 0.26 | 0.79 | 0.13 | 0.62 | 0.73 | 0.75 | 0.50 | 0.77 | 0.51 | 0.25 |
| ND DCIS (M) | | | | | | | | | | | | |
| | Cell 1 | Cell 2 | Cell 3 | Cell 4 | Cell 5 | Cell 6 | Cell 7 | Cell 8 | Cell 9 | Cell 10 | Average | Std Dev |
| PM | 35.05% | 25.15% | 19.29% | 14.52% | 17.56% | 38.57% | 40.81% | 33.41% | 29.45% | 26.89% | 28.07% | 9.02% |
| Nuc | 35.96% | 25.96% | 46.16% | 54.29% | 48.52% | 24.65% | 20.24% | 28.67% | 31.58% | 32.95% | 34.90% | 11.27% |
| Cytosolic | 38.99% | 48.90% | 34.55% | 31.19% | 33.92% | 36.77% | 38.96% | 37.92% | 38.97% | 40.16% | 38.03% | 4.75% |
| PM:Nuc | 0.97 | 0.97 | 0.42 | 0.27 | 0.36 | 1.56 | 2.02 | 1.17 | 0.93 | 0.82 | 0.95 | 0.55 |
| OC 10A (A) | | | | | | | | | | | | |
| | Cell 1 | Cell 2 | Cell 3 | Cell 4 | Cell 5 | Cell 6 | Cell 7 | Cell 8 | Cell 9 | Cell 10 | Average | Std Dev |

| | | | | | | | | | | | | |
|---|---|---|---|---|---|---|---|---|---|---|---|---|
| PM | 15.70% | 11.62% | 15.25% | 10.75% | 15.61% | 18.50% | 16.76% | 15.80% | 9.51% | 5.69% | 13.52% | 3.96% |
| Nuc | 20.68% | 35.36% | 16.32% | 13.81% | 13.80% | 22.12% | 31.12% | 34.38% | 38.94% | 22.30% | 24.88% | 9.35% |
| Cytosolic | 63.61% | 53.02% | 68.43% | 75.44% | 70.59% | 59.38% | 52.12% | 49.82% | 51.55% | 72.01% | 61.60% | 9.66% |
| PM:Nuc | 0.76 | 0.33 | 0.93 | 0.78 | 1.13 | 0.84 | 0.54 | 0.46 | 0.24 | 0.26 | 0.63 | 0.31 |
| OC 10A (M) | | | | | | | | | | | | |
| | Cell 1 | Cell 2 | Cell 3 | Cell 4 | Cell 5 | Cell 6 | Cell 7 | Cell 8 | Cell 9 | Cell 10 | Average | Std Dev |
| PM | 20.19% | 10.63% | 9.67% | 12.07% | 8.83% | 7.13% | 4.19% | 10.45% | 6.96% | 5.62% | 9.57% | 4.46% |
| Nuc | 36.09% | 30.88% | 33.75% | 33.69% | 37.51% | 21.45% | 31.01% | 19.39% | 24.79% | 29.07% | 29.76% | 6.11% |
| Cytosolic | 43.72% | 58.49% | 56.59% | 54.24% | 53.65% | 71.42% | 64.80% | 70.17% | 68.25% | 65.31% | 60.66% | 8.83% |
| PM:Nuc | 0.56 | 0.34 | 0.29 | 0.36 | 0.24 | 0.33 | 0.14 | 0.54 | 0.28 | 0.19 | 0.33 | 0.14 |
| ND 10A (A) | | | | | | | | | | | | |
| | Cell 1 | Cell 2 | Cell 3 | Cell 4 | Cell 5 | Cell 6 | Cell 7 | Cell 8 | Cell 9 | Cell 10 | Average | Std Dev |
| PM | 4.66% | 4.06% | 5.22% | 6.07% | 6.14% | 7.13% | 4.23% | 5.15% | 5.41% | 3.31% | 5.14% | 1.13% |
| Nuc | 44.15% | 76.56% | 74.52% | 76.28% | 76.42% | 78.63% | 77.41% | 78.60% | 68.82% | 71.79% | 72.32% | 10.36% |
| Cytosolic | 51.19% | 19.39% | 20.26% | 17.65% | 17.44% | 14.23% | 18.36% | 16.25% | 25.77% | 24.90% | 22.54% | 10.69% |
| PM:Nuc | 0.11 | 0.05 | 0.07 | 0.08 | 0.08 | 0.09 | 0.05 | 0.07 | 0.08 | 0.05 | 0.07 | 0.02 |
| ND 10A (M) | | | | | | | | | | | | |
| | Cell 1 | Cell 2 | Cell 3 | Cell 4 | Cell 5 | Cell 6 | Cell 7 | Cell 8 | Cell 9 | Cell 10 | Average | Std Dev |
| PM | 3.60% | 5.05% | 5.79% | 3.34% | 4.17% | 4.48% | 8.73% | 5.34% | 5.36% | 7.58% | 5.34% | 1.70% |
| Nuc | 70.19% | 65.16% | 60.58% | 69.03% | 60.84% | 31.42% | 70.37% | 73.84% | 65.15% | 61.06% | 62.76% | 11.93% |
| Cytosolic | 26.21% | 29.80% | 33.63% | 27.63% | 34.99% | 64.10% | 20.90% | 20.82% | 29.49% | 31.36% | 31.89% | 12.27% |
| PM:Nuc | 0.05 | 0.08 | 0.10 | 0.05 | 0.07 | 0.14 | 0.12 | 0.07 | 0.08 | 0.12 | 0.09 | 0.03 |

**Supplementary Table S3. Single-cell compartment fractions and PM:nuclear ratios in MCF10DCIS.com and MCF10A cells under normal-density and overconfluent conditions stained with Abcam ab39260 and MilliporeSigma ZRB3155.**

For each condition, measurements from 10 individual cells are shown for plasma-membrane (PM), nuclear, and cytoplasmic signal fractions, together with the corresponding PM:nuclear signal ratio. Mean values and standard deviations are reported for each condition-antibody pair. OC indicates overconfluent conditions, ND indicates normal-density conditions, A denotes Abcam ab39260, and M denotes MilliporeSigma ZRB3155. These data support the orthogonal antibody comparison and mechanosensing-associated PM relocalization analyses shown in **Fig. 3**.

| Stratum | Progressed / Total (PM+) | Progressed / Total (PM–) | OR (95% CI) | Fisher p | Cox HR (95% CI) | Wald p | Log-rank p |
|---|---|---|---|---|---|---|---|
| Lower grade (intermediate + low) | 14 / 18 | 2 / 8 | 10.50 (1.54–71.76) | 0.016 | 3.24 (0.85–12.41) | 0.0859 | 0.0703 |
| High grade | — | — | Not estimable (few PM–) | — | 1.85 (0.11–32.06) | 0.6723 | 0.2280 |

**Supplementary Table S4. Grade-stratified association of PM-TRPV4 with invasive progression**

Fisher's exact test (two-sided) with Woolf 95% CIs (Haldane-Anscombe correction for zero cells). Wald p-values are reported for Cox models; log-rank p-values shown for Kaplan–Meier comparisons. Lower-grade stratum = intermediate + low grade. OR not estimable for high grade due to sparse PM- counts.

| Risk Factor | Progressed | Non-Progressed | OR (95% CI) | P-Value |
|---|---|---|---|---|
| PM-TRPV4+ | 22 / 24 (91.7%) | 13 / 20 (65.0%) | **5.92 (1.07 – 32.87)** | **0.057** |
| High Grade | 8 / 24 (33.3%) | 10 / 20 (50.0%) | 0.50 (0.15 – 1.69) | 0.359 |
| ER Negative | 3 / 24 (12.5%) | 4 / 20 (20.0%) | 1.78 (0.34 – 9.16) | 0.682 |
| Cardiovascular Disease | 16 / 24 (66.7%) | 12 / 20 (60.0%) | 1.33 (0.39 – 4.58) | 0.757 |
| Smoking | 10 / 24 (41.7%) | 9 / 20 (45.0%) | 0.87 (0.26 – 2.89) | 1.000 |
| Metabolic Disease | 9 / 24 (37.5%) | 6 / 20 (30.0%) | 1.40 (0.40 – 4.96) | 0.752 |

**Supplementary Table S5. Association of Clinical and Pathologic Factors with DCIS Progression to Invasive Ductal Carcinoma (n = 44)**

Univariable associations between clinicopathologic factors and invasive progression from ductal carcinoma in situ (DCIS). Progressed and non-progressed groups represent counts and percentages of patients positive for each risk factor. Odds ratios (OR) and 95% confidence intervals (CI) were calculated using two-sided Fisher's exact tests, with CIs derived by the Woolf logit method. Only plasma-membrane TRPV4 positivity demonstrated a strong association with progression, whereas histologic grade, ER status, and comorbidities were not predictive. Bold indicates $p < 0.10$.

| | **Intensive** | **Lumpectomy alone** |
|---|---|---|
| PM-TRPV4+ | 28 / 34 (82.4%) | 6 / 9 (66.7%) |
| PM-TRPV4− | 6 / 34 (17.6%) | 3 / 9 (33.3%) |

OR = 2.33 (95% CI 0.45–12.06); Fisher p = 0.367.  N = 43

**Supplementary Table S6. PM-TRPV4 status vs surgery group (Fisher's exact)**

Associations of surgical management and disease site with PM-TRPV4 and progression. Two-by-two Fisher's exact tests report odds ratios (OR), 95% confidence intervals (CI; Woolf logit with Haldane–Anscombe correction for zeros), and p-values for PM-TRPV4 status vs surgery group (treatment selection)

| | **Intensive** | **Lumpectomy alone** |
|---|---|---|
| Progression | 17 / 34 (50.0%) | 6 / 9 (66.7%) |
| No progression | 17 / 34 (50.0%) | 3 / 9 (33.3%) |

OR = 0.50 (95% CI 0.11–2.33); Fisher p = 0.467. N = 43

**Supplementary Table S7. Progression vs surgery group (Fisher's exact)**

Associations of surgical management and disease site with PM-TRPV4 and progression. Two-by-two Fisher's exact tests report odds ratios (OR), 95% confidence intervals (CI; Woolf logit with Haldane–Anscombe correction for zeros), and p-values for progression vs surgery group (outcome association). Intensive treatment includes lumpectomy with adjuvant radiation or mastectomy; Lumpectomy alone indicates no radiation/mastectomy recorded.

| Disease Site | n |
|---|---|
| Ipsilateral | 13 |
| Contralateral | 10 |
| Unknown | 1 |

**Supplementary Table S8. Disease site among progressed cases**

Distribution of disease site (ipsilateral vs contralateral) among progressed cases.

| | PM+ | PM− |
|---|---|---|
| Ipsilateral | 11 / 22 (50.0%) | 2 / 2 (100.0%) |
| Contralateral | 11 / 22 (50.0%) | 0 / 2 (0.0%) |

OR = 0.20 (95% CI 0.00–0.87); Fisher p = 0.482.  N = 24

**Supplementary Table S9. Laterality (Ipsilateral vs Contralateral) by PM among progressed (Fisher's exact)**

Among progressed cases, Fisher OR for ipsilateral (vs contralateral) events by PM-TRPV4 status. Analyses are two-sided with α=0.05.
